# Altered brain network topology in children with Auditory Processing Disorder: a resting-state multi-echo fMRI study

**DOI:** 10.1101/2022.04.05.22273478

**Authors:** Ashkan Alvand, Abin Kuruvilla-Mathew, Ian J. Kirk, Reece P. Roberts, Mangor Pedersen, Suzanne C. Purdy

## Abstract

Children with auditory processing disorder (APD) experience hearing difficulties, particularly in the presence of competing sounds, despite having normal audiograms. There is considerable debate on whether APD symptoms originate from bottom-up (e.g., auditory sensory processing) and/or top-down processing (e.g., cognitive, language, memory). A related issue is that little is known about whether functional brain network topology is altered in APD. Therefore, we used resting-state functional magnetic resonance imaging data to investigate the functional brain network organization of 57 children from 8 to 13 years old, diagnosed with APD (n=28) and without hearing difficulties (healthy control, HC; n=29). We applied complex network analysis using graph theory to assess the whole-brain integration and segregation of functional networks and brain hub architecture. Our results showed children with APD and HC have similar global network properties and modular organization. Still, the APD group showed different hub architecture. At the nodal level, we observed decreased participation coefficient (PC) in auditory cortical regions in APD, including bilateral superior temporal gyrus and left middle temporal gyrus. Beyond auditory regions, PC was also decreased in APD in bilateral posterior temporo-occipital cortices, left intraparietal sulcus, and right posterior insular cortex. Correlation analysis suggested a positive association between PC in the left parahippocampal gyrus and the listening-in-spatialized-noise-sentences task where APD children were engaged in auditory perception. In conclusion, our findings provide evidence of altered brain network organization in children with APD, specific to auditory networks, and shed new light on the neural systems underlying children’s listening difficulties.

## 1. INTRODUCTION

Auditory Processing Disorder (APD) is an umbrella term for listening difficulties that result from a deficit in the neural processing of auditory stimuli or speech (Keith et al., 2019; Dawes & Bishop, 2010; American Speech-Language-Hearing Association (ASHA), 2005; Dillon & Cameron, 2021). It is estimated that 5.1% of school-aged children have difficulties understanding speech in competing background noise such as the classroom despite having no hearing loss based on their pure tone audiogram (Hind et al., 2011; Purdy et al., 2018). Keith et al. (2019) estimated that in New Zealand, where this study is conducted, the prevalence of APD in school-aged children is around 6.2% or higher. Classroom difficulties of children with APD include difficulty hearing in background noise, poor sound localization, inconsistency in answering questions, frequent requests for repetition, trouble understanding and complying with verbal instructions, and poor attention (American Academy of Audiology (AAA), 2010; de Wit et al., 2016; Chermak et al., 2002). Some of these children also show deficits in their speech and language skills, including reading and writing (Sharma et al., 2009; Dawes & Bishop, 2010; Barker et al., 2017; Gokula et al., 2019). In the past 40 years, children with these symptoms have been invited for further specialized testing at audiology clinics to investigate the possibility of APD (AAA, 2010). Although there is no global consensus amongst audiologists and speech-language pathologists regarding diagnosing developmental APD (Moore et al., 2013), substantial efforts have been made to develop clinical guidelines for the assessment and treatment of children with APD (ASHA, 2005; Keith et al., 2019; Wilson, 2018; Iliadou et al., 2018).

Complex auditory processing happens at all levels within the auditory system; the ability to localize, discriminate, recognize auditory patterns, or discriminate temporal sound features could be impaired (Wilson, 2018; ASHA, 1996). This impairment in auditory perception is characterized as caused by deficiencies or developmental differences in the central auditory nervous system (CANS), through which auditory signals are transmitted via the cochlear nerves to the auditory cortices (British Society of Audiology (BSA), 2011; AAA, 2010; ASHA, 1996). Neural encoding of auditory signals is associated with complex, parallel and serial processing within auditory regions in CANS and other processing in the higher-order cortical regions (AAA, 2010; Moore, 2012; Ponton et al., 1996).

Due to the heterogeneity of behaviors and symptoms observed in children with APD (e.g., many presents with comorbid memory and attention deficits, for example), many different cortical areas, such as superior temporal, inferior parietal and inferior frontal areas involved in higher-order language and cognitive functions have been implicated (Moore, 2012, 2015; Poremba et al., 2003; Fritz et al., 2010). Symptoms of APD overlap with other sensory or cognitive neurodevelopmental disorders (Sharma et al., 2009; Dawes & Bishop, 2010; Moore et al., 2013; AAA, 2010). APD can co-occur with reading and language deficits in children with diagnoses of specific language impairment (SLI), reading disorder/dyslexia, autism spectrum disorder (ASD). However, this does not occur for all children (Jerger & Musiek, 2000; Sharma et al., 2009; Halliday et al., 2017; Mealings & Cameron, 2019; Dawes et al., 2009). Additionally, children with APD can display attention (Moore et al., 2010; Gyldenkærne et al., 2014) and/or memory problems (Sharma et al. 2009; 2014), but many cases of APD who do not have memory or attention deficits. Thus, the overlap of cognitive, language and hearing difficulties in children with APD and other neurodevelopmental disorders is controversial for researchers who have debated whether auditory sensory processing deficits cause the listening difficulties in children diagnosed with APD (bottom-up approach: related to the ear or CANS) or cognitive deficits (top-down approach: related to cognitive function deriving from multi-modal processing) (Moore, 2012; Moore & Hunter, 2013; McFarland & Cacace, 2014; Dillon et al., 2012; Cacace & McFarland, 2013). According to the BSA (2011) position statement, “APD is characterized by a poor perception of both speech and non-speech sounds” (p.4) and “attention is a key element of auditory processing, and that poor attention makes a major contribution to APD” (p.6). Other clinical guidelines (ASHA, 2005; AAA, 2010) do not specify higher-order cognitive, communicative and language-related functions in their statements as contributing factors for listening difficulties in APD (de Wit et al., 2016). In this debate, Cacace and McFarland (2013) asserted that APD could not be considered a distinct disorder if modality specificity could not be demonstrated with certainty. Additionally, Moore and Hunter (2013) suggested APD could be distinguished from cognitive deficits based on modality specificity. Hence, the complex interactions between cognitive, language, reading, and auditory processing abilities in the brain of children with APD are not entirely understood. The potential for neuroimaging research to address this controversy and help understand of the top-down processing mechanisms in children with APD has been increasingly recognized (Moore & Hunter, 2013; Bartel-Friedrich et al., 2010; AAA, 2010; Pluta et al., 2014; Stewart et al., 2021).

In the last decade, links between atypical functional connectivity in large-scale brain organization and neurodevelopmental disorders, such as autism, Tourette’s syndrome, or attention deficit hyperactivity disorder (ADHD), have been identified (Zhang & Raichle, 2010; Openneer et al., 2020; Cocchi et al., 2012; Li et al., 2014; Sadeghi et al., 2017; Faridi et al., in press). Resting-state functional Magnetic Resonance Imaging (rsfMRI) can be utilized to explore the functionally important aspects of whole-brain intrinsic networks without requiring the participant to perform a task (van den Heuvel & Hulshoff Pol, 2010), and hence this has been a fast-growing technique in brain imaging to study neurological brain disorders in at-risk developmental populations (Fornito & Bullmore, 2010; Fornito et al., 2013; Rosazza & Minati, 2011). Using rsfMRI data, functional connectivity can be estimated between anatomically distributed regions by recording temporal correlations of spontaneous fluctuations in the Blood-Oxygenation-Level-Dependent (BOLD) signal (Biswal et al., 1997). In the APD literature, to our knowledge, there have been only three resting-state fMRI studies that investigated this intrinsic activity (Pluta et al. 2014; Stewart et al., 2021, Preprint; Hoyda et al., 2021, Preprint). The earliest of these studies investigated 13 children with diagnosed APD (without other neurological signs) and 15 healthy control (HC) children (Pluta et al., 2014) using regional homogeneity (ReHo) to investigate changes in the pattern of the default mode network (DMN). Intrinsic connectivity within this network has been associated with attentional impairment (Bonnelle et al., 2011). The ReHo results indicated decreased functional activity in the superior frontal gyrus and posterior cingulate cortex/precuneus (Pluta et al., 2014); these areas are involved in control and attention and implicate the role of DMN in these cognitive processes (Castellanos et al., 2008). Pluta et al. (2014) argued that modality-specific perceptual dysfunction could not differentiate between children with neurodevelopmental APD and those with attention deficits. More recent research involving 81 children (n=42 Listening difficulties/APD; n=39 typically developing/TD) used a region of interest (ROIs) approach to assess how speech perception and listening networks (Phonology, Intelligibility, Semantics) are different in children with APD compared to TD (Stewart et al., 2021). Stewart and colleagues (2021). These networks were based on differential activation of areas during contrasting listening and language processing tasks. Children with APD had increased functional connectivity in the left inferior frontal gyrus (Broca’s area) and left posterior middle temporal gyrus; these regions are implicated in language production and comprehension and lexical and semantic processes (Hagoort, 2014; Acheson et al., 2013; Hickok & Poeppel, 2004). Stewart and colleagues (2021) also reported that in the Semantics network, children with APD had stronger functional connectivity in the right parahippocampal gyrus compared to auditory areas such as left Heschl’s gyrus, left middle temporal gyrus (MTG), right superior temporal gyrus (STG), and right planum temporale. Still, they had weaker connections in the left temporal fusiform cortex and right superior temporal gyrus (Stewart et al., 2021). Stewart et al. (2021) concluded that atypical neurological aspects of APD are connected with language comprehension, consistent with the high rate of language disorder comorbidity in children with APD (Sharma et al., 2009; Dawes & Bishop, 2010; Barker et al., 2017). A follow-up study by the same research team investigating speech and higher-order functioning in the same group of children with APD (Hoyda et al., 2021) defined ROIs for speech perception, speech production, language comprehension, naming and executive networks based on the Neurosynths database (Yarkoni et al., 2011), and then looked for group differences in pairwise functional connectivity. Their results showed significantly decreased connectivity in the APD group compared to TD within the executive function network in the left caudate and left mid frontal gyrus suggesting a link between executive function and APD (Hoyda et al., 2021). These studies (Pluta et al., 2014; Stewart et al., 2021; Hoyda et al., 2021) have investigated functional connectivity in children with APD based on predefined ROIs, but it is not clear yet how these ROIs and their interaction could contribute to altered brain network connectivity in APD. Thus, despite growth in the application of rsfMRI in neurodevelopmental research, the functional topological organization of children with APD remains largely unknown.

Recently, graph theoretical approaches have been more widely used to analyze rsfMRI data to measure the macroscopic structural and functional attributes of brain networks (Bullmore & Sporns, 2009; Power et al., 2011). This approach, also known as network neuroscience, offers a mathematical framework for investigating the local and global properties of neural systems (Bassett & Sporns, 2017; Sporns, 2011), and is considered a promising tool for understanding brain networks and their behaviors (Fornito et al., 2016). The human brain can be conceptualized as a complex network topology (Watts & Strogatz, 1998; Sporns, 2011), which is optimally balanced in segregation (local connectivity) and integration (global connectivity) of information flow (Sporns, 2013). These complex topologies can be represented as multiple nodes (brain regions) representing local functioning and a smaller set of nodes illustrating global functioning (Bullmore & Sporns, 2009). Brain organization displays scale-free, small-world (van den Heuvel et al, 2008; Bassett & Bullmore, 2006; Fornito et al., 2016), hierarchical modularity (Meunier et al., 2010), hub (Power et al., 2013), and rich-club (van den Heuvel & Sporns, 2011) architectures. Hub architecture consists of important brain regions that interact with many other regions, facilitate functional integration, and play a key role in optimal information flow (Rubinov & Sporns, 2010). This crucial importance of hubs makes them vulnerable spots in brain networks; an alteration in brain hubs is one of the most consistent findings in network-based studies (Meunier et al., 2009; Rubinov & Bullmore, 2013; Crossley et al., 2014; Roger et al., 2020). Thus investigating brain hub organization can provide new insights into the neural mechanisms underlying APD and may reveal differences in brain topologies.

To the best of our knowledge, this is the first rsfMRI study that has applied graph theoretical approaches to understand the large-scale brain organization of children with APD. The present study used rsfMRI and the complex network analysis method to examine the whole brain functional topology in children diagnosed with APD and children without complaints of listening difficulties (healthy controls, HC). This research generated functional brain connectomes to measure global and brain hub topology and examine brain network differences between APD and HC. Studies of children with developmental disorders generally do not show significant differences in all global properties of the brain (Armstrong et al., 2016; Sadeghi et al., 2017; Chen et al., 2019; Zhang et al., 2019), and hence more insights may be gained by examining regional brain topology. Based on these previous studies of children showing similarities in their global brain network organization, we hypothesized that the APD and HC groups would not differ in their resting-state functional connectivity or global topological architecture (i.e., whole-brain averaged network topology). However, children with APD showed atypical regional brain topology, specifically in cortical temporal regions associated with auditory function and hub organization.

## 2. METHODS

Approval for this study was granted by The University of Auckland Human Participants Ethics committee (Date: 18/10/2019, Ref. 023546). Before the start of the study, children and their parents signed assent and consent forms as per the requirement of the Ethics committee.

### 2.1 Participants

A total of 66 children aged between 8 -14 years participated in this study, but only 57 participants remained for the analysis; nine children were excluded from the further data analysis due to excessive head motions (*n* = 4), incidental findings (*n* = 1), and uncompleted scans (*n* = 4). Among the 57 included participants, 28 children were already diagnosed with APD (13 boys, M_age_= 10.92, range 8.58-13.41), and 29 children were healthy controls (14 boys, M_age_=11.91, range 9.75-14.08). All children with APD were recruited from SoundSkills clinic in Auckland, New Zealand (https://soundskills.co.nz). They were diagnosed formally with APD according to New Zealand guidelines for standardized testing APD test battery (Keith et al., 2019). Children in the HC control group were recruited via posted flyers and online advertisements. They were excluded if diagnosed with hearing loss, learning difficulties, and any neuropsychiatric conditions or taking medication known to affect the central nervous system. Four children in the HC group (14%) were not experiencing any learning difficulties but had diagnoses of ASD (n=1), ADHD (n=2), or dyslexia (n=1). Twelve children with APD were also diagnosed with other comorbid developmental disorders such as dyslexia (n=8), ADHD (n=1), attention deficit disorder (ADD, n=2) and developmental language disorder (DLD, n=1). The comorbidities were allowed as these disorders coexist with APD (Dawes & Bishop 2010; O’Connor, 2012; Sharma et al., 2009).

### 2.2 Procedure

Children and their parents were invited to take part in two separate sessions on a single day to complete the hearing tests and the MRI scan. Most participants completed the entire procedure on the same day. All testing was conducted at the University of Auckland’s Clinics and the Centre for Advanced MRI (CAMRI), located at the Grafton Campus, Auckland, New Zealand.

#### 2.2.1 Session 1: Hearing assessment

In this session, all participants underwent 30 minutes of hearing tests, including otoscopy, pure tone air conduction audiometry (PTA), and tympanometry to check hearing acuity and screen for middle ear disease and atypical ipsilateral middle ear muscle reflexes (Roup et al., 1998). Otoscopy results for all children were normal. PTA thresholds for every participant were no more than 20 dB HL at the octave frequencies from 0.25 to 8 kHz in each ear, and tympanogram results in both ears were indicative of normal middle ear function (static admittance in range 0.2 to 1.6 mmho, peak pressure between -100 and +20 daPa). All children were also tested on the listening-in-spatialized-noise-sentences (LiSN-S; (Cameron & Dillon, 2007; 2008) test, which is a tool for assessing the ability of individuals to understand sentences in the presence of competing speech presented from different directions and using the same or different talkers. LiSN-S scores are presented as z-scores compared to normative findings for a child of the same age (Cameron et al., 2011). The low-cue scores (speech reception threshold, SRT) reflect listening skills when no spatial or vocal cues are available to the listener to distinguish target sentences from the distractor stories. High-cue SRT scores assess listening when both vocal and spatial cues are available. Talker, Spatial, and Total Advantage scores are difference scores that reflect listeners’ ability to use differences in either the voices of the speakers, the physical location of the target and distractors, or both of these types of cues, respectively, to identify the sentences in the presence of competing speech. The advantage scores are difference scores that control for variations in overall speech perception abilities across participants and accurately reflect auditory processing within the central auditory system. Specific details of the LiSN-S test are explained in Besser et al. (2015).

#### 2.2.2 Session 2: MRI scan

##### Participant preparation

To prepare the children for the MRI scan, a few different strategies were employed to reduce anxiety and movement during the scan session, as described in Wilke et al. (2018). Firstly, before the appointment, all participants’ parents or caregivers were encouraged to video clips to familiarize with the MRI scan procedure. Secondly, children were instructed about all the procedures using a miniature Lego scanner during their hearing session (Fig. 1); this helped reduce children’s concerns about the scanning session. Finally, all children and their parents or caregivers were invited to attend a 20 min preparation session in the mock scanner before the actual scan started. In this session, the instructions were explained again to the children to know what would happen in the actual scan session to have a successful session. Children were instructed and practised how to stay still during the scan by playing a statue game in the mock scanner and showing them pictures illustrating how head motion can cause blurry brain images. To reduce the anxiety of children, parents or caregivers also were allowed to have the option to stay in the scan room with their children where they could see them - only a few parents chose this option. Children were allowed to bring their non-magnetic toys to the scan room. All participants were asked to use the restroom immediately before the scan as recommended by Wilke et al., (2018).

**Figure 1.**
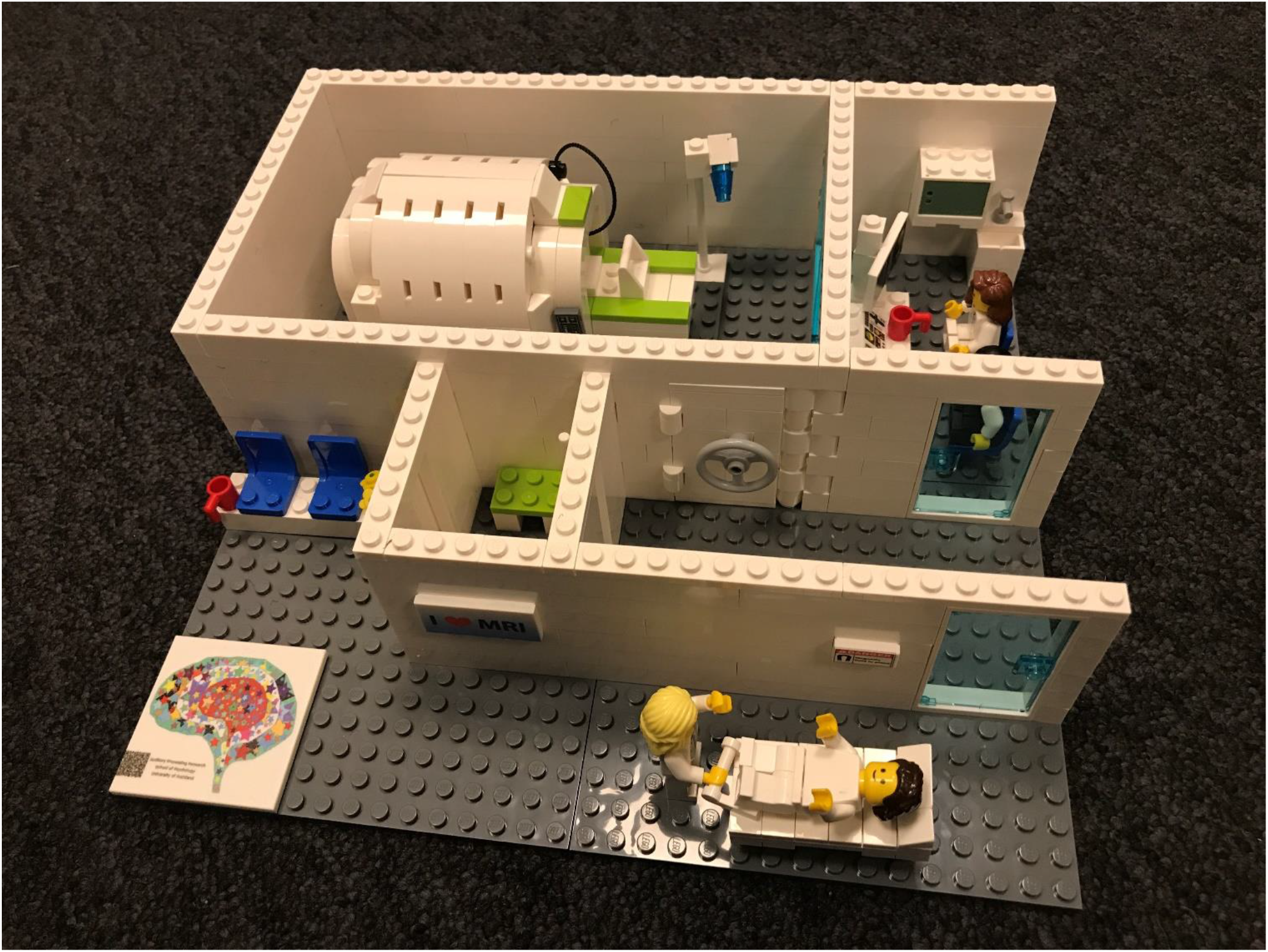
A picture of a miniature scanner model provided for this study from the Amazings team (https://www.amazings.eu/p/mri.html).

##### MRI scan

This study utilized a 20-channel head coil for recording brain activity in all sequences, which is larger than the widely used 32-channel head coil. The 20-channel head coil was more comfortable for the children and allowed better control of movement using padding to fixate the head (Greene et al., 2016) whilst accommodating the headphones needed for accessible communication. During the scan, children were asked to lay still, not fall asleep, and keep their eyes open. Children were given earplugs and headphones to minimize the loudness of the MRI noise. For the initial T1-image sequence (5 min), children were required to watch an animation of their choice to keep them entertained, while for the rsfMRI sequence (7 min; 20 s), they were asked to stay still and stare at the cross presented on the screen. Children’s heads were stabilized with cushions to diminish head movements. At the end of the rsfMRI sequence, participants were asked to rate how awake they felt during the scan from 1 to 10, where 1 represents falling asleep and 10 is completely awake.

### 2.3 MRI data acquisition

MRI data were acquired on a 3T Siemens MAGNETOM Skyra scanner (Siemens, Erlangen, Germany). The high-resolution structural T1-image was acquired for co-registration using a magnetization-prepared rapid acquisition gradient echo (MPRAGE) sequence with 1 mm isotropic resolution (Field of view (FOV) = 256 mm, 208 sagittal slices in a single slab, TR = 2000 ms, Echo time (TE) = 2.85 ms, Flip angle = 8 degrees, slice thickness =1mm, acquisition time 286 sec).

rsfMRI data were acquired using multi-echo/multiband (ME/MB) echo-planar imaging (EPI) sequences, customized for the pediatric population (TR 1700 ms, TEs 15, 31.63, 48.26 ms, flip angle 83 deg, multi-band factor 2, GRAPPA PAT mode, 3.2 mm isotropic voxels, 3 mm slice thickness, Field of View 202 mm, and 46 slices with full brain coverage) these sequences were adapted from Marusak et al.’s (2018) study. The Siemens embedded system automatically discards the first 10 time points of the BOLD data to achieve equilibrium, resulting in 250 volumes for the analysis. As stated in the literature, MB EPI improves temporal resolution (Preibisch et al., 2015; Demetriou et al., 2018), and using the ME approach reduces signal dropout in brain regions and improves the temporal signal to noise ratio (tSNR) (Kundu et al., 2012; 2015).

### 2.4 Image preprocessing

All anatomical and functional images were converted to Nifti file sets using dcm2niix software (version 11/11/2020) developed by Rorden Lab (Rorden et al., 2007) (https://github.com/rordenlab/dcm2niix), and all Nifti files were structured according to Brain Imaging Data structure (BIDS v1.8.2, Gorgolewski et al., 2017) (Fig. 2A).

**Figure 2.**
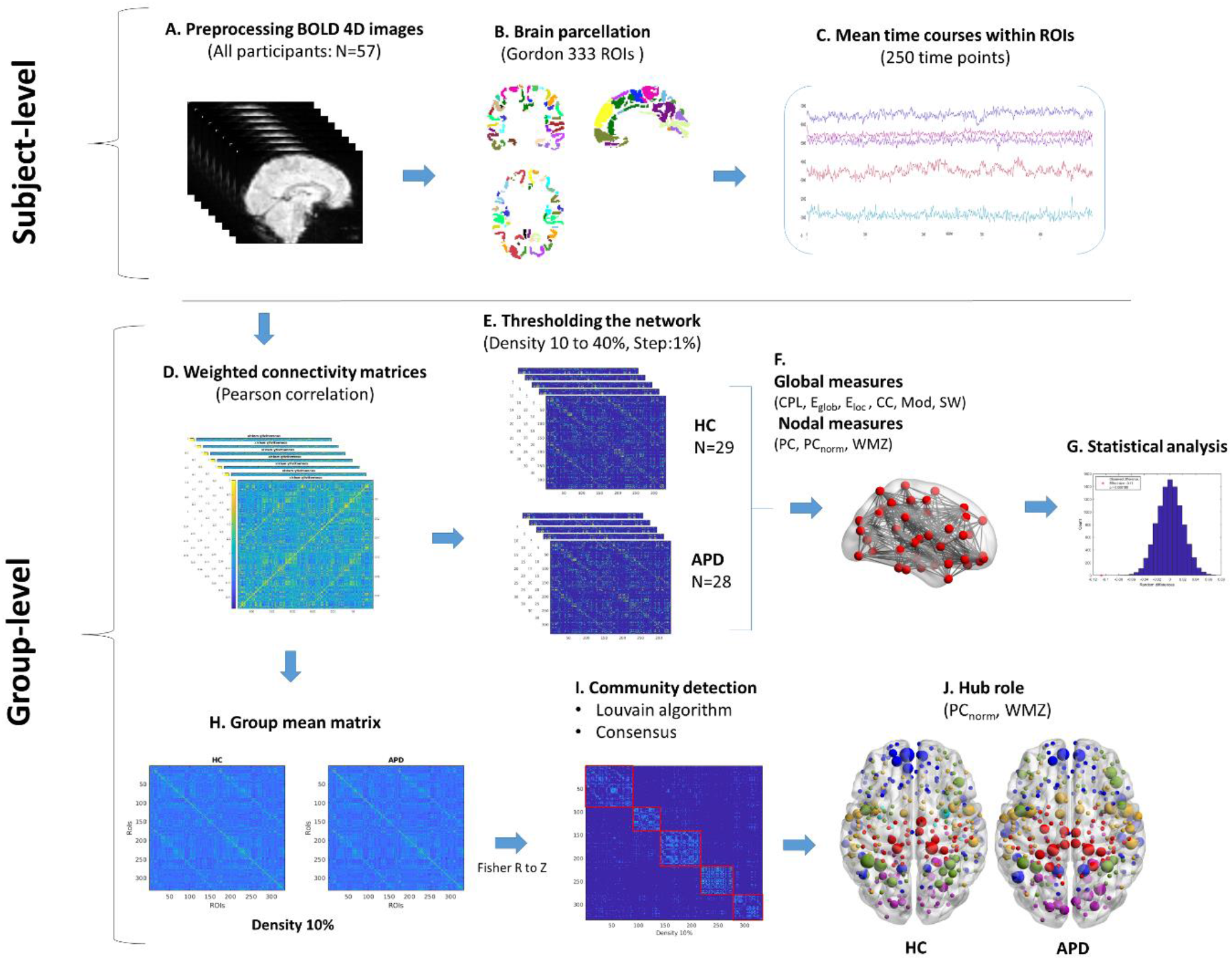
Schematic representations of study pipeline. First, all participants’ raw functional data were preprocessed and denoised (A) and then parcellated into 333 ROIs using Gordon parcellation (B), corresponding time series were extracted and averaged within each ROI to calculate individual connectivity matrices (D), afterwards, each connectivity matrix was thresholded and graph theory of global and nodal measures were calculated (F) and then area under the curve at sparsity of 10 to 40% was computed for statistical analysis with the permutation test (G). Also, the group averaged matrices were calculated for each group at the network density of 10% (H) and using the Louvain algorithm modular architecture of each group was revealed (I) to identify hubs and their functional roles based on WMZ and PC_norm_ metric (J).

#### Quality control

The functional and anatomical data quality was assessed using MRIQC’s visual reports (Esteban et al., 2017). All data were visually checked for correct alignment to the corresponding T1w image and signal artifacts. Data were inspected for head motions based on quality control measures, including carpet plots (Power, 2017), framewise displacement (FD), DIVARS (D referring to the temporal derivative of time courses, VARS referring to RMS variance over voxels) (Power et al., 2012; 2014). Participants were excluded under a stringent regime based on these criteria: mean FD greater than 0.25 mm; or FDs greater than 5 mm (Parkes et al., 2018).

#### fMRIPrep workflow

Preprocessing of resting-state BOLD data and high-resolution T1-weighted (T1w) structural data was performed according to the fMRIPrep v20.2.3 pipeline (Esteban et al., 2019), which is based on Nipype 1.6.1 (Gorgolewski et al., 2011). The following description is based on the boilerplate published by the fMRIPrep pipeline, and it is covered by a “no rights reserved” (CC0) license. Internal operations of the fMRIPrep pipeline utilize a combination of software, including FSL v5.0.9 (Smith et al., 2004), ANTs v2.3.3 (Avants et al., 2009), Freesurfer v6.0.1 (Dale et al., 1999), AFNI v16.2.07 (Cox, 1996), and Nilearn v0.6.2 (Abraham et al., 2014).

Each T1w image was corrected for intensity non-uniformity (INU) with N4BiasFieldCorrection (Tustison et al., 2010), distributed with ANTs, and used as T1w-reference throughout the workflow. The T1w-reference was then skull-stripped with a Nipype implementation of the antsBrainExtraction.sh workflow, using the target template. Brain tissue segmentation of cerebrospinal fluid (CSF), white matter (WM), and grey matter (GM) was performed on the brain-extracted T1w using FSL’s fast (Zhang et al., 2001). Volume-based spatial normalization to MNIPediatricAsym:cohort-4: res-2 (MNI’s unbiased standard MRI template for pediatric data from the 7.5 to 13.5 age range) was performed through nonlinear registration with antsRegistration (ANTs), using brain-extracted versions of both T1w reference and the T1w template.

For each child’s functional data, first, all three echoes were slice-time corrected using AFNI’s 3dTshift (Cox & Hyde, 1997). Head-motion parameters for the BOLD reference are estimated using FSL’s mcflirt (Jenkinson et al., 2002). A B0-nonuniformity map (or field map) was also estimated based on a phase-difference map calculated with a dual-echo GRE (gradient-recall echo) sequence. The field map was then co-registered to the target EPI reference run and converted to a displacements field map with FSL’s Fugue and co-registered to the T1w reference with nine degrees of freedom using FSL’s FLIRT(FMRIB’s Linear Image Registration Tool) (Jenkinson & Smith, 2001) with the boundary-based registration (Greve & Fischl, 2009) cost-function. The BOLD time-series were resampled onto their original, native space by applying a single, composite transform to correct for head-motion and susceptibility distortions. Then, the T2* map was estimated according to Posse et al.’s (1999) method and used to optimally combine BOLD data across echoes using tedana’s t2smap (Kundu et al., 2012; 2015). The BOLD time-series were then resampled into standard space (MNIPediatricAsym:cohort-4:res-2). Automatic removal of motion artifacts using independent component analysis (ICA-AROMA, Pruim et al., 2015) was performed on the functional data’s time series. As a result, the “aggressive” noise-regressors were collected and placed in the corresponding confounds file (*_desc-confounds_timeseries.tsv). Several confounding time-series were calculated based on the preprocessed BOLD: FD (Jenkinson et al., 2002; Power et al., 2014), DVARS, and three region-wise global signals. FD and DVARS are calculated for each functional run, both using their implementations in Nipype (Gorgolewski et al., 2011). The three global signals are extracted within the CSF, the WM, and the whole-brain masks. For more details about the fMRIPrep pipeline, see https://fmriprep.org/en/latest/workflows.html.

#### Denoising strategy

The unsmoothed output from fMRIPrep labeled by the suffix _space-MNIPediatricAsym_cohort-4_res-2_desc-preproc_bold.nii.gz was used for further processing. First, spatially normalized BOLD time courses were linearly detrended, and then intensity normalization was applied to mode 1000 units. Then identified time series from WM, CSF, whole-brain global signals, and noise components identified by ICA-AROMA were regressed from the BOLD data by the fsl_regfilt function with an aggressive denoising strategy (ICA-AROMA+8P+4GSR; Parkes et al., 2018). ICA-AROMA (Pruim et al., 2015) utilizes FSL’s MELODIC tool (Beckmann et al., 2005) for decomposing BOLD data into spatially independent components (ICs) to categorize ICs as BOLD or non-BOLD signals. The advantage of using ICA-AROMA as described in the literature (Parkes et al., 2018; Ciric et al., 2017) is that this is the most effective pipeline for mitigating motions related artifacts and reducing spurious connectivity (Power et al., 2012; Satterthwaite et al., 2012; Van Dijk et al., 2012). It is also worth noting that the combination of multi-echo fMRI and ICA-AROMA can improve the quality of BOLD signals extracted from components and the functional connectivity between each pair of ROIs (Dipasquale et al., 2017). Finally, the residual denoised BOLD data were bandpass filtered between 0.008 and 0.08 Hz using a Fourier transform (Parkes et al., 2018). They were spatially smoothed with a 6 mm FWHM (full-width half-maximum) kernel. Furthermore, as Parkes and colleagues (2018) suggested, quality control functional connectivity benchmarks were also calculated to measure the efficacy of different denoising strategies. The results are presented in the Supplementary Materials (See Fig. S1).

### 2.5 Brain network construction

#### Defining nodes and edges

A network is composed of nodes and edges where each node represents brain regions in a network, and edges represent the connections between all brain regions. In this study, nodes are defined based on anatomically or functionally parcellated cortical brain regions, and edges are estimated by statistical interdependence (i.e., correlations) in BOLD signals among pairwise brain regions (Friston et al., 1993). The number of nodes in the estimation of the brain network varies considerably for different parcellations methods. Because the application of different parcellations may result in different outcomes from the network analysis (Zalesky et al., 2010a), we used two different functional parcellations (i.e., Gordon and Schaefer parcellations) for defining nodes of the brain network (Gordon et al., 2016; Schaefer et al., 2018). These parcellation masks were used because of their accuracy in defining spatial GM boundaries, their neurobiological plausibility in the analysis of rsfMRI data (Hallquist & Hillary, 2019; Eickhoff et al., 2018) and their utilization in other network neuroscience studies (Hagler et al., 2019; Feczko et al., 2018; Faskowitz et al., 2020). For each participant, preprocessed rsfMRI data were parcellated in sets of 333 ROIs produced from a published data-driven parcellation method (Gordon et al., 2016) (See Fig. 2B). These 333 cortical regions were represented as nodes in the current study’s network topology. For validation, the functional data were also parcellated into 300 separated ROIs (Schaefer et al., 2018). This data-driven parcellation includes 300 brain regions defined based on 17 functionally parcellated networks from Yeo et al. (2011) study. Subsequently, the time series of all voxels within each region were averaged (Fig. 2C). Next, Pearson’s correlation *r* was computed based on the BOLD signal between all brain regions to determine pairwise functional connectivity strength. This resulted in a subject-specific symmetric and undirected weighted connectivity matrix of size 333×333. Each pair-wise element represents Pearson’s correlation between the mean time courses between brain regions (Fig. 2D). Networks are typically thresholded (removal of non-significant network edges) and binarized, but this process is semi-arbitrary (van den Heuvel et al., 2017). Studies have shown that the density and organization of brain networks vary between subjects, which may lead to systematic changes in results depending on the threshold chosen (Wen et al., 2011). Based on these methodological limitations, we use a matching approach of thresholding by using a range of network sparsity thresholds to ensure all subject-specific networks contain the same number of nodes *and* edges (Zhang et al., 2011; Achard & Bullmore, 2007; Langer et al., 2013). We computed connectivity matrices with a network density ranging from 1 to 40% (with a 1% increment) to investigate the topological properties of the brain network. For the analysis, a minimum bound of 10% was selected to prevent graph fragmentation at a sparser threshold and an upper bound of 40% was chosen due to its liberal estimate of cerebral connectivity and its neurobiological plausibility for brain functional organization (Fornito et al., 2010; Zhang et al., 2011) (Fig. 2E). It also ensured that all brain networks indicated network properties of small-worldness for all densities (Fig. S2 C).

### 2.6 Graph theory analysis

For investigating the topological aspects of the functional network, graph metrics of each individual’s network were calculated on the undirected weighted functional network. The graph theory computation was carried out by functions from the Brain Connectivity Toolbox (BCT v 03/03/2019, https://sites.google.com/site/bctnet/) in MATLAB R2019b (https://mathworks.com/).

#### Network integration and segregation (whole-brain averaged measures)

Functional integration is defined as combining specialised information across nodes and shows how the network can share the information in the distributed nodes (Rubinov & Sporns, 2010). Measures of characteristic path length (CPL) and global efficiency (E_glob_) were calculated to estimate network integration. A short path length indicates that each node can reach other nodes with few steps (i.e., the minimum number of edges connecting two connected brain regions), or a path composed of few edges; the average of the shortest path between all pairs of nodes is defined as CPL (Rubinov & Sporns, 2010). E_glob_ is also a measure that indicates the capacity of a network for transferring parallel information in each pair of nodes, and it is defined as the inverse of the shortest path length between nodes (Latora & Marchiori, 2001). Therefore CPL and E_glob_ are primary metrics to measure network integration (Achard & Bullmore, 2007). Functional segregation refers to the ability in the brain that specialized processing happens in the densely interconnected group of brain regions (Rubinov & Sporns, 2010). To estimate the local properties of brain networks, a range of metrics such as mean local efficiency (E_loc_), Clustering Coefficient (CC), and Modularity Optimization (Mod) were calculated. E_loc_ metric measures the network capability in transferring the information at the local level (node’s neighborhood). It is defined as the average inverse shortest path between two nodes (Rubinov & Sporns 2010; Latora & Marchiori, 2001). CC is defined as the fraction of triangular connected nodes around the node of interest and indicates the degree to which neighbors tend to cluster with each other in the network (i.e., *Cliquiness*; Watts & Strogatz, 1998), hence the mean CC is considered as a direct measure of brain segregation (Rubinov & Sporns 2010). Mod is a segregation measure that indicates the presence of densely interconnected nodes (i.e., modules) and indicates the size and composition of these modular structures (Girvan & Newman, 2002; Rubinov & Sporns, 2010). Small-worldness (SW) is an analogy based on the small-world phenomenon (Watts & Strogatz, 1998), indicating that a biological system such as the brain has a structure that is neither regular or random (Latora & Marchiori, 2001). This property is highly clustered like a regular lattice network but has a similar characteristic path length to a random network (Watts & Strogatz, 1998). Hence, the SW metric can present the segregation and integration of the network and shows a high global and local efficiency of a complex system (Pievani et al., 2011). To examine the SW property of each individual’s brain network, 100 random networks (BCT, *randomio_und* function) with the same topological properties of empirical networks and a wiring cost of 20 were generated based on the Maslov-Sneppen Null network model that preserves the degree distribution of the original networks (Maslov & Sneppen, 2002) (See Fig. 2F).

#### Hub detection

Important nodes or hubs participate in many interactions within a network and play a key role in optimal information flow in the brain networks (Rubinov & Sporns 2010; Stam, 2014; Power et al., 2013). As proposed by Guimerà and Nunes Amaral (2005), hubs and their topological roles can be quantified by measures of within-module degree z-score (WMZ) and participation coefficient (PC). WMZ is a measure that estimates how strongly a node is connected to its module relative to other nodes within the same community (intra-modular connections). PC is an inter-modular degree that describes if nodes are distributed uniformly across many modules or are concentrated within a module. This metric reveals the modular segregation and intermodular integration of the network in the form of the provincial hub (nodes with high WMZ, low PC) and connector hubs (nodes with high WMZ and high PC) (Rubinov & Sporns, 2010; Fornito et al., 2016; Guimerà & Nunes Amaral, 2005; Meunier et al., 2009). To perform hub detection, first, group average matrices were calculated for each group at the top 10% of the connections because of the high rank of their correlation strength (Alavash et al., 2019) (Fig. 2H). Afterwards, the Louvain community detection algorithm (BCT: community_louvain.m) was applied to reveal the modular brain structure (Blondel et al., 2008) due to its fast and reliable procedure in detecting communities (Betzel, 2020).

Since this algorithm is heuristic and stochastic, this process was repeated 1000 times to reach a consensus for determining community labels (concencus_und.m, Betzel, 2020) (Fig. 2I). Once all nodes were assigned to their modules, WMZ (module_degree_zscore.m) and PC were calculated. Recent work by Pedersen and colleagues (2020) has shown that to reduce the influence of intra-modular connectivity, which may result in inaccurate inference in a network with different modular sizes and also maximize the identification of interconnected nodes, it is beneficial to normalize PC with randomly generated null networks (PC_norm_; Pedersen et al., 2020). Hence in this study PC_norm_ was computed by *participation_coef_norm*.*m* (https://github.com/omidvarnia/Dynamic_brain_connectivity_analysis/) function based on 1000 randomizations which were previously shown as an adequate iteration for estimating stable PC_norm_ (Pedersen et al., 2020) (See Fig. 2J). Lastly, brain hubs and their role in networks were identified based on the criteria described according to an earlier study by Meunier and colleagues (2009) as follows: connector hub: WMZ>1 and PC_norm_ >0.5; provincial hub: WMZ>1 and PC_norm_ <0.5.

### 2.7 Statistical analysis

Group characteristics and behavioral variables were tested with two-sample t-tests to analyze differences in age, head motion (mean FD), and LiSN-S results. Pearson chi-square tests were also used to compare handedness and gender distribution between groups.

#### Differences in edge-wise functional connectivity

To assess the group differences across the entire connectome, we used the network-based statistics toolbox (NBS, https://sites.google.com/site/bctnet/comparison/nbs) (Zalesky et al., 2010b). The NBS is a connectome-wide analysis approach that improves statistical power over common mass-univariate correction methods such as FDR by localizing the strength of connectivity to identify significant effects at the components level (sub-networks) (Zalesky et al., 2010b; Fornito et al., 2016). In NBS, these subnetworks are constructed by applying an initial threshold to the data (p<0.05, uncorrected). The observed subnetwork sizes were then compared with the empirical null distribution of maximal component sizes acquired by the permutation test (10000 times) (Fornito et al., 2016). This evaluation of observed component sizes with regards to the null distribution of maximal sizes controls for the Family-wise error (FWE) for resulting inference on subnetworks (Fornito et al., 2016). First, all individuals’ connectivity matrices were Fisher’s R to Z transformed (Mudholkar, 2014). Two-sample t-tests were then performed for each pairwise connection linking 333 brain regions to test group differences in functional connectivity in either direction (two-tailed hypothesis test). Age was included as a covariate for these analyses.

#### Differences in hub measures

To assess whether there are group differences in hub detection measures of PC, PC_norm_, and WMZ, the area under the curve (AUC) across the sparsity range of 10 to 40% was calculated. The AUC was chosen because integrating network density costs could control the monotonic transformation of a weight set of the weighted graph (Ginestet et al., 2011) and it may improve the sensitivity in detecting the case-control group differences (Achard & Bullmore, 2007; Hosseini et al., 2012; van den Heuvel et al., 2017). Two-sample t-tests assuming unequal variances between APD and HC groups were carried out on the AUC of each network measure in the Permutation Analysis of Linear model software (PALM; https://fsl.fmrib.ox.ac.uk/fsl/fslwiki/PALM/UserGuide). PALM uses a permutation test in a general linear model framework while controlling for false-positive errors under weak or unreasonable assumptions. The data are primarily exchangeable under the null hypothesis (Winkler, Ridgway, Webster, Smith, Nichols., 2014). To test the null hypothesis that the group means were all equal, for each network measure, all values randomly were assigned to all subjects and the mean differences were recomputed between two randomized groups. The randomization was repeated 20000 times, and the 95% confidence interval was calculated and used as the critical value of significance testing (p<0.05). The effect of age as a nuisance confound was also controlled during the randomization (demeaned). To control the multiple comparisons problem, the p-value of all network measures was corrected using false discovery rate (FDR) correction using an alpha level of 0.05 (Benjamini & Hochberg, 1995) (See Fig. 2G).

#### Relationship between hub measures and LiSN-S variables

Exploratory partial correlation analysis was performed to assess the association between nodal measures of PC, PC_norm_ and WMZ with LiSN-S results (z-scored) using a permutation test (20000 randomizations) and controlling the effect of age (*p* values corrected with FDR correction, *p*<0.05). For this analysis, three participants (HC=1, APD=2) were excluded due to missing behavioral data for correlation analysis between the brain and LiSN-S data.

#### Meta-analytic interpretation

Neurosynth (Yarkoni et al., 2011), a data-driven tool for functional meta-analysis, was utilized for data interpretation. The Neurosynth database contains more than 14000 functional neuroimage studies that allow the identification of associated cognitive terms with the brain activation pattern. Neurosynth extracted and visualised the top 20 cognitive terms (excluding redundant, anatomical terms) associated with significant ROIs as a word cloud in MATLAB R2019b. After identifying ROIs showing significant group differences, the Neurosynth database was searched for these ROIs to identify relevant cognitive terms. For each significant region, up to 20 associated cognitive terms were selected and visualized for each ROI based on their meta-analytic co-activation scores (Pearson r, uncorrected).

## 3. RESULTS

### 3.1 Demographics and spatialized listening scores

Group comparisons of demographic variables, including motion (mean FD), handedness, and gender, showed no significant differences. However, APD and HC were significantly different in age (HC>APD). Group comparisons of LiSN-S scores only indicated significant differences in Talker Advantage scores (HC>APD), although Fig. 3 shows poorer advantage scores on average for the APD group. Further demographic details are depicted in Table 1.

**Figure 3.**
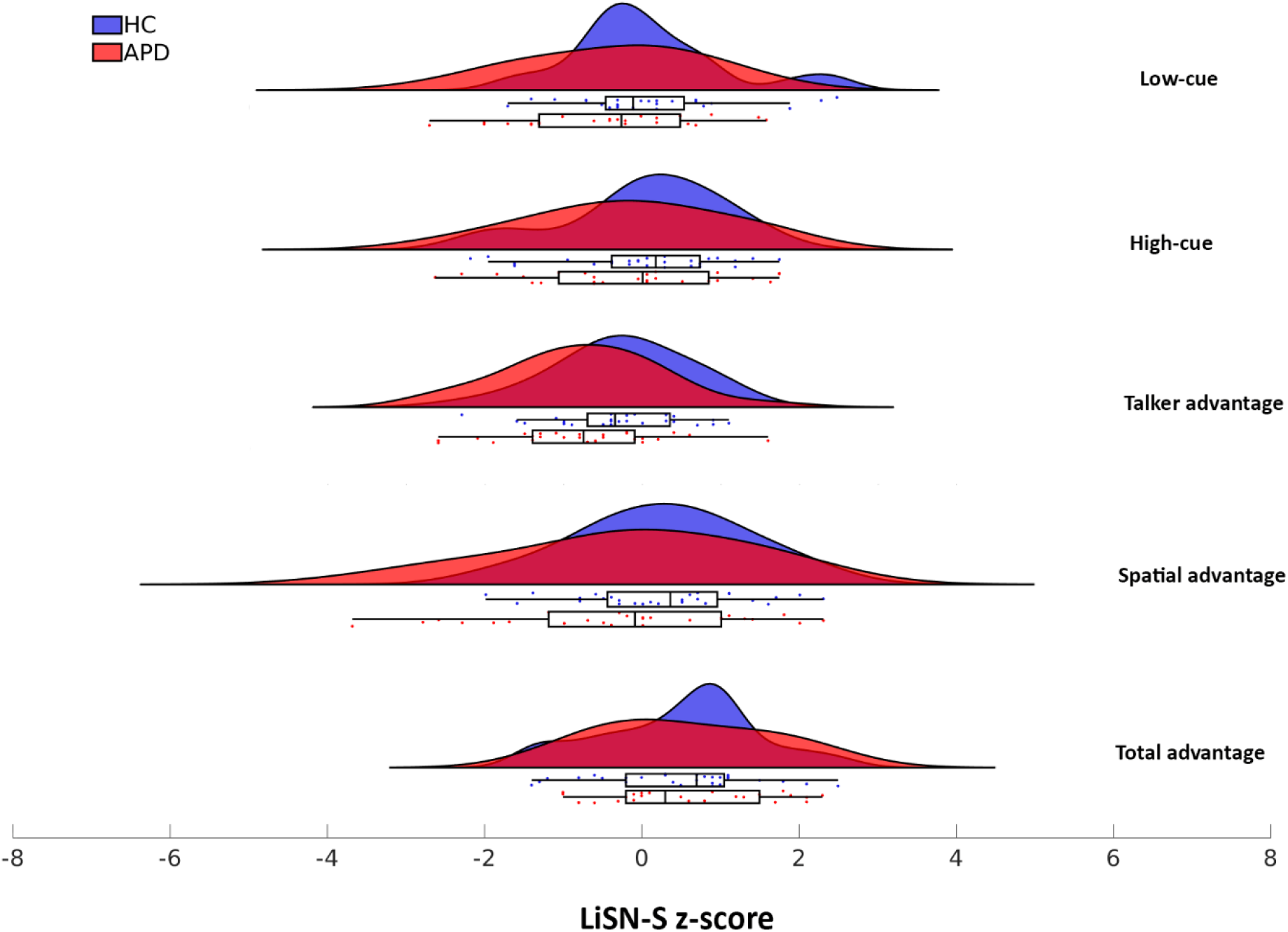
Distribution of LiSN-S z-scores (Total advantage, Spatial advantage, Talker advantage, High-cue, Low-cue) between APD and HC groups

**Table 1.** Demographic and group characteristics

|  | APD (n=28) | HC (n=29) | Test Statistic | p value |
| --- | --- | --- | --- | --- |
| Age (years) | 10.92±1.55 | 11.91±1.39 | 2.546<sup>a</sup> | 0.014 |
| Gender (male/female) | 13/15 | 14/15 | 0.020<sup>b</sup> | 0.889 |
| Handedness (right/left) | 23/5 | 26/2 | 1.631<sup>b</sup> | 0.202 |
| Mean FD (mm) | 0.10±0.03 | 0.11±0.04 | 0.962<sup>a</sup> | 0.340 |
| LiSN-S | (n=26) | (n=28) |  |  |
| Total advantage | 0.53±1.04 | 0.47±0.98 | -0.216<sup>a</sup> | 0.830 |
| Spatial advantage | -0.25±1.56 | 0.25±1.06 | 1.398<sup>a</sup> | 0.168 |
| Talker advantage | -0.76±0.98 | -0.25±0.82 | 2.052<sup>a</sup> | 0.045 |
| High cue | 0.28±1.08 | 0.48±0.89 | 0.746<sup>a</sup> | 0.459 |
| Low cue | -0.35±1.12 | 0.07±0.98 | 1.507<sup>a</sup> | 0.138 |

**Note:** Data are presented as n or mean ± standard deviation. HC, healthy control; APD, auditory processing disorder; FD, framewise displacement; LiSN-S, Listening in Spatialized Noise -Sentences Test. Between-group differences were tested with two-sample t-tests<sup>a</sup> and Pearson chi-square tests<sup>b</sup>

### 3.2 Edge-wise functional connectivity

NBS assessed group differences in functional connectivity. The results showed no significant difference in edge-wise connectivity between APD and HC groups after controlling for multiple comparisons (FWR-corrected).

### 3.3 Whole-brain averaged network measures

Investigating network segregation indicated higher mean CC and mean E_loc_ in the APD group than HC. However, group comparisons revealed no statistical differences across the range of network density thresholds (1 to 40%). Fig. S2 presents results from Gordon parcellation with 333 functionally separated regions. Outcomes from other global measures are provided in supplementary materials.

### 3.4 Module and hub roles in APD topological network

Examining the modular organization of HC and APD groups (Density 10%), there were similarities (Fig. 4) but also distinct differences regarding the size and topological role of community structures between groups. The modular community detection analysis revealed five functional modules in both groups, including default mode-ventral attention (Fig. 4 Module I, Blue), somatomotor (Fig. 4 Module II, Red), limbic (Fig. 4 Module III, Orange), visual (Fig.4 Module IV, Purple), and frontoparietal-dorsal attention (Fig. 4 Module V, Green) (Yeo et al., 2011). The most extensive module in both groups was default mode-ventral attention, with 86 ROIs in APD and 90 ROIs in HC groups, comprising the default mode (DM), cinguloparietal, and ventral attention regions. The smallest module in both groups was somatomotor with 50 and 38 ROIs, respectively (See Table 2). Fig. 5 displays brain hubs and their role in the network (i.e., connector or provincial) based on WMZ and PC_norm_. A major difference between the two groups was driven by the somatomotor module, where APD had 10 provincial hubs compared to HC with 4 provincial hubs. Also, the APD group had several regions where nodes were identified as hubs, in contrast to HC where these regions were not hubs, in the DM (ROIs #6, #162), somatomotor-hand (ROIs #2, #50, #57, #163), and dorsal attention (DA; ROI #74) regions. More details about group differences in hub regions are presented in Table S1.

**Figure 4.**
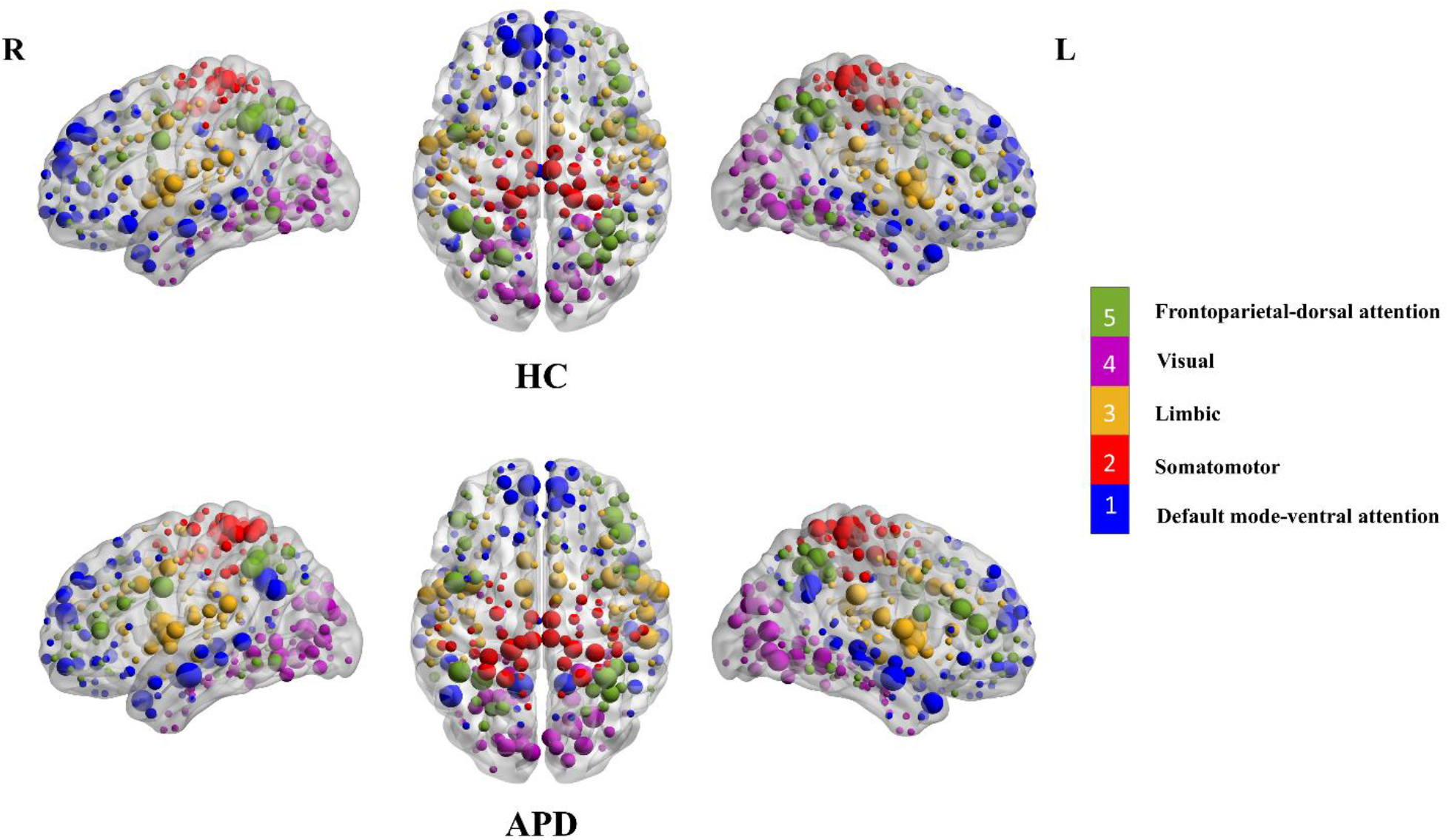
Modular organization of functional brain network in APD and HC groups based on their group averaged matrices (density 10%) and Gordon parcellation. All 333 ROIs are marked by different colors (each color represents a distinct module) and then mapped into cortical templates in sagittal and axial view using BrainNet Viewer (Xia et al., 2013). Larger nodes indicated hubs widely interconnected in the module. Abbreviation: L - left hemisphere, R - right hemisphere.

**Table 2.**
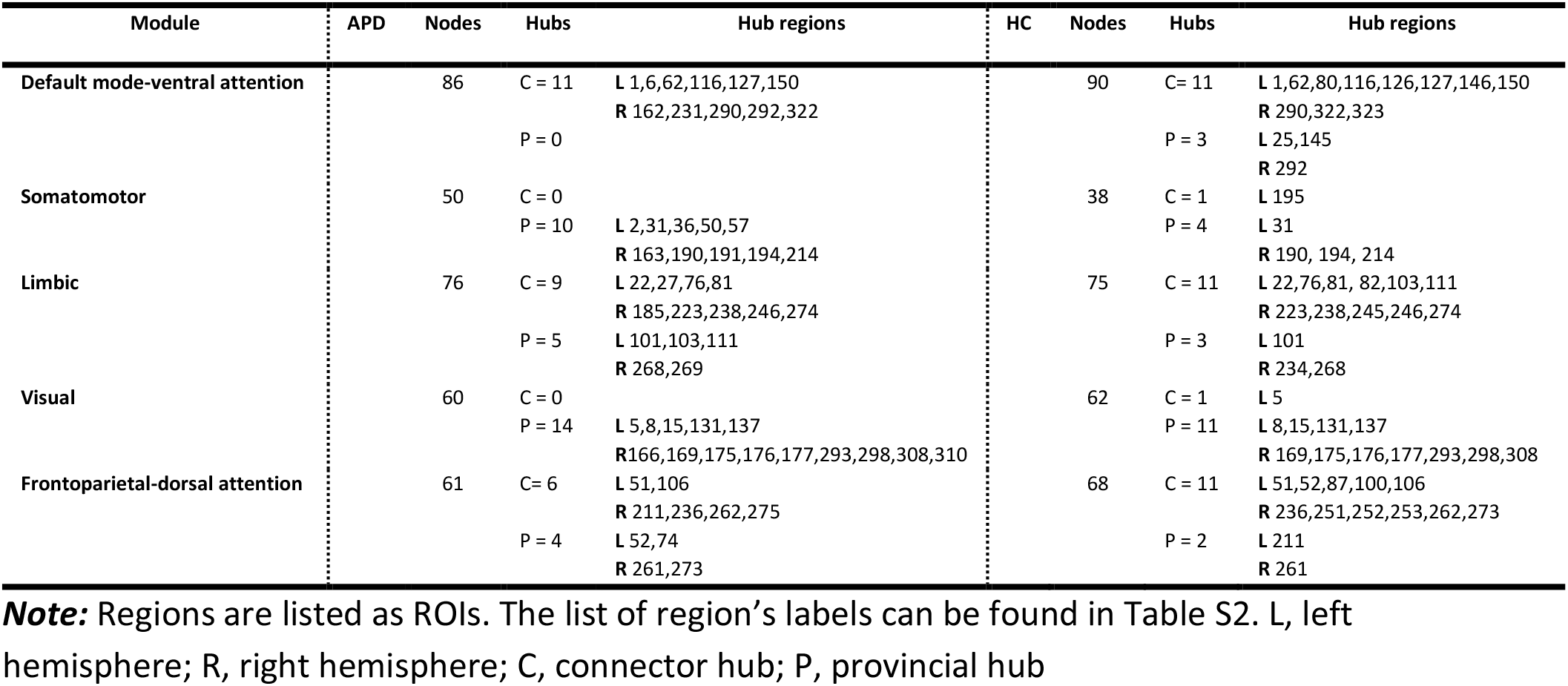
Summary of modular properties and identified hub regions

**Figure 5.**
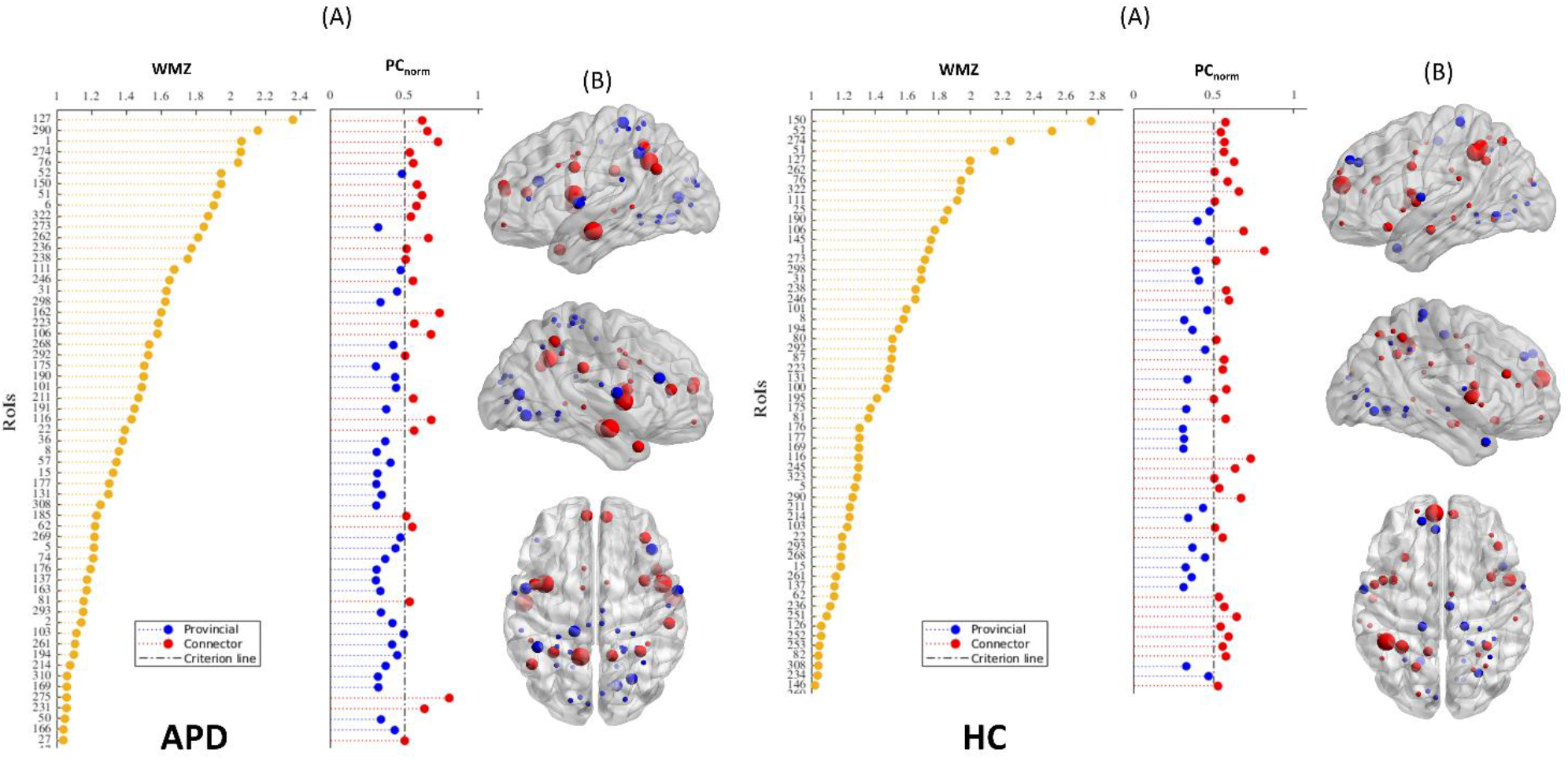
Brain hubs and their roles in APD and HC groups. (A): Within module degree z-score (WMZ) and normalized participation coefficient (PC_norm_) are shown for regional nodes in both groups. Nodes with high WMZ (WMZ>1, yellow dots) are considered to be hubs. Hub nodes with high PC (PC>0.5, red dots) are connector hubs and those with low PC_norm_ (PC_norm_<0.5, blue dots) are provincial hubs. (B): Corresponding connector and provincial hubs are also presented on the brain surfaces in sagittal (Left and Right) and axial views constructed by BrainNet Viewer.

Results from the group analysis based on the nodal measure of PC revealed a significant group difference in the region of DMN. This was observed in the right STG in node #331 (DMN, *p*=0.0333, t=3.985, FDR corrected). Fig. 6 illustrates the result for the PC measure (AUC 10-40%) of 333 functionally separated brain regions. The red color indicates an elevation of the PC metric in the HC group (APD<HC). A similar analysis was carried out based on the Schaefer parcellation (17 functional networks, 300 ROIs) to assess the robustness of this result. The outcome from this analysis exhibited differences in the following networks (All *p*<0.05, FDR-corrected): left and right STG (ROIs #147, #294, respectively), left MTG (ROI #124), left and right posterior temporo-occipital (pTO; #56, #298), left intraparietal sulcus (IPS; ROI #93) as well as right posterior insular cortex (INS; ROI #216). There is an overlap of regions #331 and #294 from the Gordon and Schaefer parcellations, respectively. More details regarding the results from the Schaefer parcellation are provided in Table S2. No significant differences between the groups were observed in the nodal measures of PC_norm_ and WMZ using the Schaefer parcellation.

**Figure 6.**
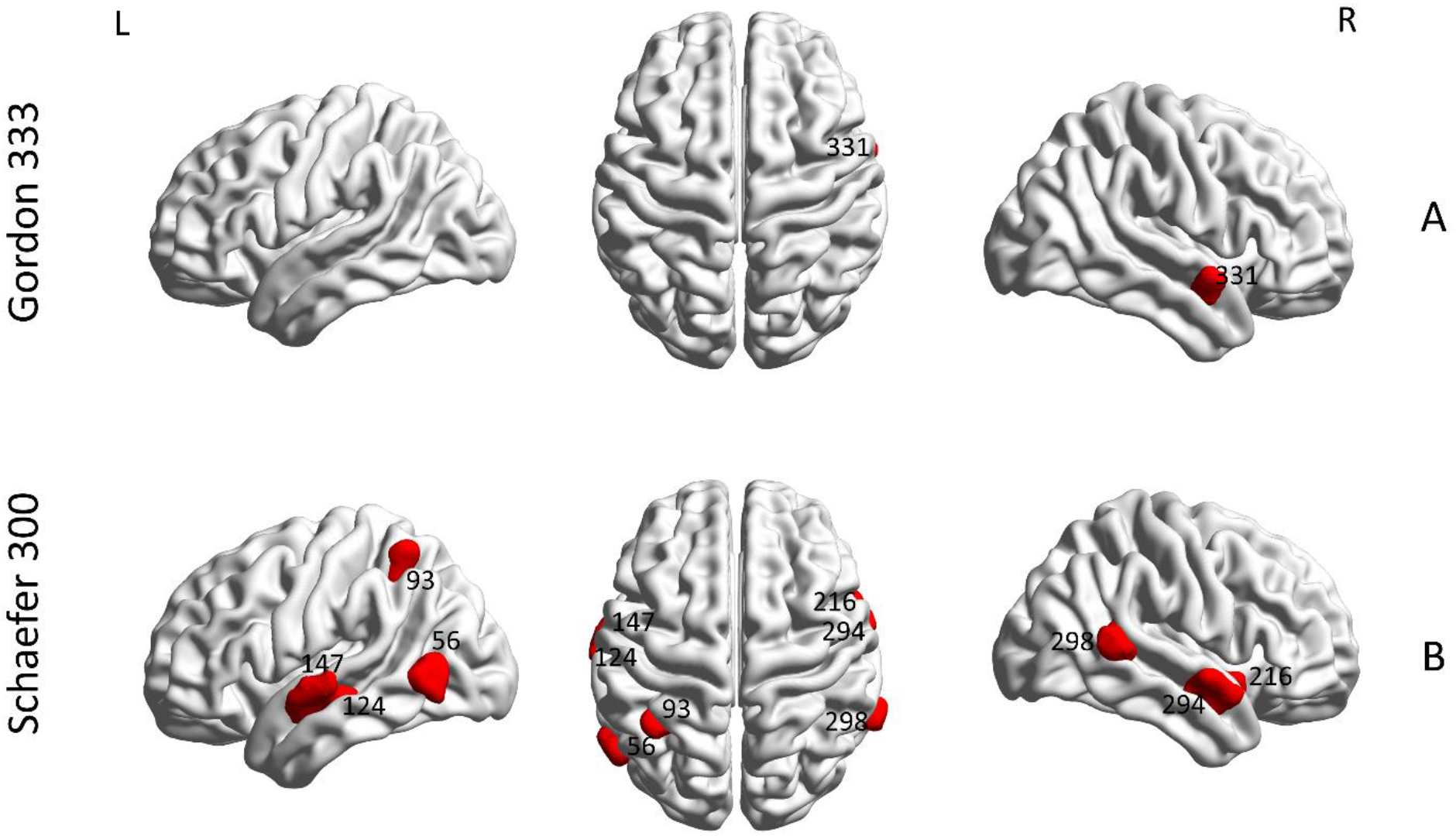
Regional differences in the PC measure between APD and HC participants (10-40% density). (A) indicates the significant difference based on the Gordon parcellation (333 ROIs) in area 331 (*p*<0.05, FDR corrected), whereas (B) shows the significant differences based on the Shaefer parcellation (300 ROIs) in the following areas: 56, 93, 124, 147, 216, 294 and 298 (*p*<0.05, FDR corrected). The group differences shown in red depict APD<HC. Images are created in BrainNet Viewer. L, Left hemisphere; R, right hemisphere.

### 3.5 Association between brain hub measures and LiSN-S variables

Partial correlation analysis indicated a significant positive correlation between PC measure (AUC 10-40%) and LiSN-S Spatial advantage in the APD group (*p*<0.05, FDR corrected). This association is observed in the left retrosplenial-temporal (RT) network (parahippocampal gyrus) in area 130 based on the Gordon parcellation (*p*=0167, Pearson r= 0.671, FDR corrected). The scatterplot and brain surface visualization of the relationship between PC and the Spatial advantage score are shown in Fig. 7. There was no significant correlation between other LiSN-S scores and PC, PC_norm_ and WMZ measures in either group.

**Figure 7.**
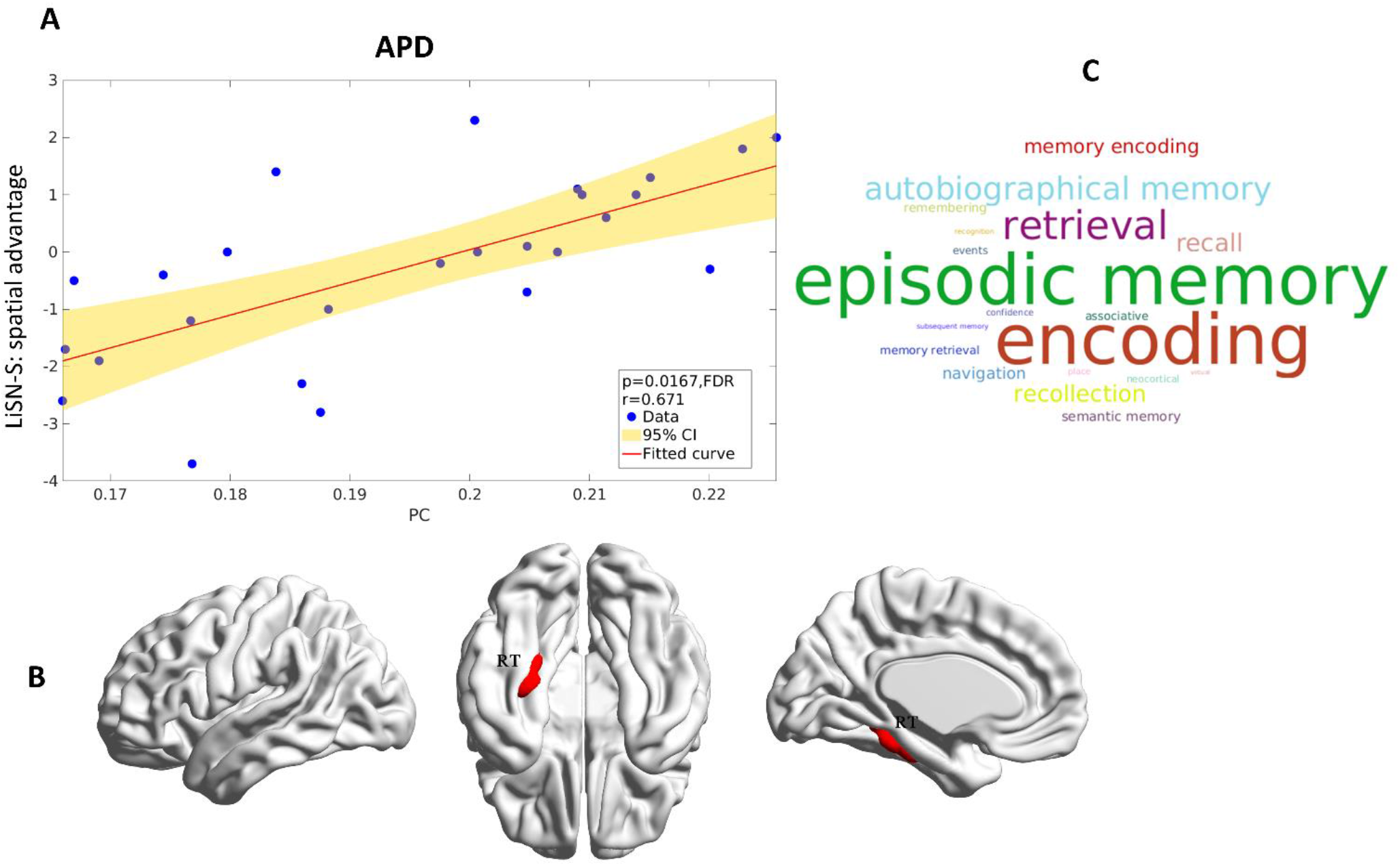
Significant association between PC measure and LiSN-S score (Spatial advantage) in auditory processing disorder (APD) group. (A) The scatter plot presents this relationship for 26 participants in the APD group; the solid line represents a regression fit (Fitted curve) to the data, and the shaded area indicates two-sided 95% confidence intervals (CI) visualized by the GRETNA plot (Wang et al., 2015). (B) Brain Surfaces showing parahippocampal gyrus part of retrosplenial-temporal network (RT; ROI #130) in the Gordon parcellation presented in sagittal, medial and ventral views created by BrainNet Viewer (Xia et al., 2013). (C) Word cloud of cognitive terms associated with RT region obtained from Neurosynth database (Yarkoni et al., 2011). The size of each cognitive term corresponds to the association of meta-analytical maps created by Neurosynth.

## 4. DISCUSSION

This is, to our knowledge, the first study to investigate the topological brain network function of children diagnosed with APD by using rsfMRI. In line with our hypothesis, our findings suggest that children diagnosed with APD demonstrate similar global network topology (integration and segregation) and edge-based connectivity (network-based statistics), to HC. However, we observed significantly decreased between-module connectivity in APD compared to HC (using the PC metric) in cortical auditory brain areas and nodes within the default mode network. Furthermore, partial correlation revealed a positive correlation between the LiSN-S behavioral measures of Spatial advantage and between-module connectivity of the parahippocampal gyrus.

### 4.1 Hub organization in APD

Network hubs are topologically more central than other brain regions. They are highly interconnected within and between networks, playing an influential role in brain network functions (van den Heuvel & Sporns, 2013). An essential characteristic of a healthy brain is the rich club of interconnected hubs; depending on the location of hubs in the brain community, hubs can have different connectivity ranges (van den Heuvel & Sporns, 2011). Hubs within a community or module are only connected to regions in that module (provincial hubs), and those connecting multiple modules are highly connected outside of the communities (connector hubs). Brain hubs are typically vulnerable in neurological disorders (Stam, 2014; Power et al., 2013; Crossley et al., 2014; Fornito et al., 2015). In the current study the default mode-ventral attention module was the most densely interconnected component in the brain across children diagnosed with APD and typically developing children. Within the default mode-ventral attention module, all hub regions in the APD group were connector hubs, however, the HC group had several provincial hubs based on this analysis. The number of connector hubs in the frontoparietal-dorsal attention module differed in the APD group compared to HC.

Previously research suggests that DMN and Frontoparietal network (FPN) are part of control-default and cross-control connector hubs subsystems (Gordon et al., 2018; 2020) and these connector hubs allow flexible control of cognition and behaviors (Bertolero et al., 2017; Gratton et al., 2018). The FPN is also believed to apply top-down regulatory control over DMN and lower-level systems via connector hubs (Dosenbach et al., 2008; Marek & Dosenbach, 2018; Gratton et al., 2018; Cole et al., 2013). The relationship between the DMN and language networks has been investigated to determine interactions of linguistic processing with internally-oriented functions of DMN (Power et al., 2011; Gordon et al., 2020). Hub differences in the APD group could suggest the shift in specific circuits between subnetworks in DMN and FPN and language networks; these subnetworks were reported by Gordon and colleagues (2020) in healthy young adults. A recent review paper by Oldham and colleagues (2021) has shown that, in the human brain, connections between hubs start to form from the developmental stage. This formation continues until brain maturation (Oldham et al., 2021; Oldham, & Fornito 2019). Our results revealed differences in the number of provincial hubs in the SM, DA, and cinguluopercular networks in the APD group compared to HC individuals. The presence of more provincial hubs in the APD group, who were aged 8 to 13 years, suggests that differences in hub localization and segregation within a module could emerge early in childhood.

### 4.2 Regional group differences in PC

Our regional hub analysis found that children with APD had less PC (between-module connectivity) than HC in the right STG (Figure 6 - ROI #331). To investigate the relationship between the right STG region and its role in cognitive functioning, we utilized the Neurosynth meta-analytic tool. The complete list of associated cognitive areas is illustrated in Fig. 8 A, B in a word cloud form. The role of the right STG in Listening, Speech and Music perception, processing of sound, and language comprehension is well established (Moerel et al., 2012; Adank et al., 2015; Trost et al., 2012; Klein & Zatorre 2011; Kriegstein & Giraud 2004; Beaucousin et al., 2007). Additionally, our complimentary results based on the Schaefer cortical atlas (Fig. 6 and Fig. 8B) were aligned with the Gordon parcellation. These findings showed not only an alteration in the right STG (ROI #294) but also indicated bilateral cortical dysconnectivity in the Left STG (ROI #147), and left MTG, (ROI #124). These regions in the bilateral supratemporal regions form part of the auditory cortex, supporting hearing and speech processing (Zatorre et al., 2002; Pandya, 1995). In our research, the bilateral auditory cortices including STG were identified as provincial non-hub (PC_norm_<0.5, WMZ<1; provincial hub is a hub with greater intra-modular connectivity) regions in DMN and were categorized as part of the default mode-ventral attention module in both groups. Human brain processing of speech and music occurs predominantly in the left and right auditory regions (Tervaniemi & Hugdahl, 2003; Zatorre et al., 2002). Speech perception is associated with activity in both left and right auditory cortices, however, left auditory regions are more specialized for phonemic and language processing (Binder et al., 2000; Binder et al., 2008; Katz, 2016). Poor phonological processing and difficulty discriminating rapid spectro-temporal characteristics of phonemes are deficits in children with APD and dyslexia (Keith et al., 2019; Burns, 2013). Stewart and colleagues (2021), using contrasting sentences involving intonation, phonetics, prosody and intelligibility in their task-based fMRI, found that linguistically meaningful units activated bilateral MTG/STG and other auditory areas (Stewart et al., 2021; Moore et al., 2020). Group differences were found for semantic and intelligibility networks. In their rsfMRI ROI-ROI analysis Hoyda and colleagues (2021) also found decreased functional connectivity bilaterally and within the left hemisphere in STG region.

**Figure 8.**
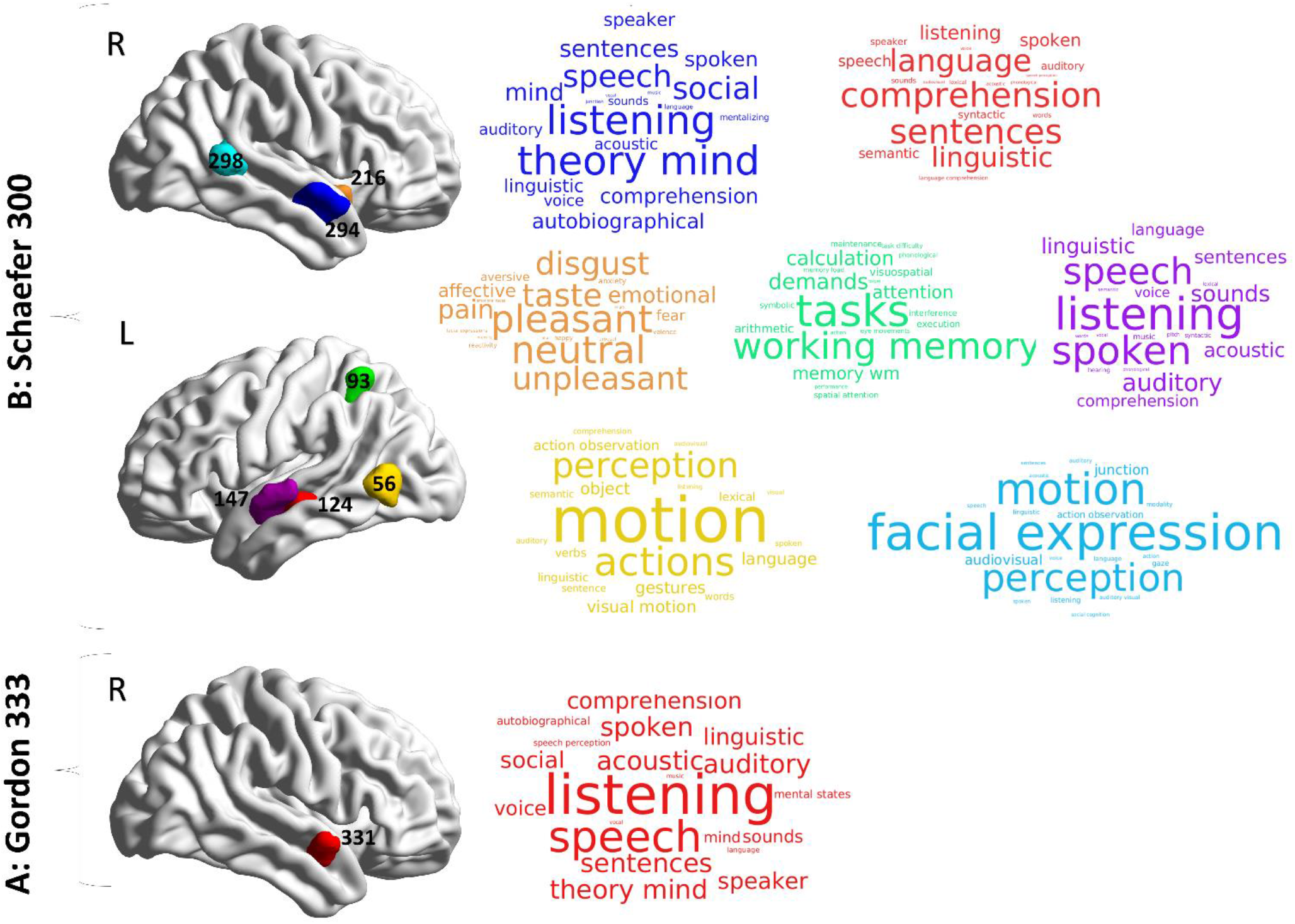
Association of meta-analytic topics with regional group differences in the PC measure. The surface templates of between-group differences in PC metrics are depicted based on Gordon (333 ROIs) (A) and Schaefer (300 ROIs) parcellations (B). Cortical regions and their association with meta-analytic terms obtained from the Neurosynth database are shown in word clouds. Each word cloud contains the top 20 relevant cognitive terms across a wide range of terms (1334 terms). Colors correspond to a different brain region and the size of each word indicates the frequency of reports in the literature (Meta-analytic coactivation score). More details about the selection criteria for cognitive terms are shown in Table S3. L: left hemisphere, R: right hemisphere.

DMN is primarily localized in the medial prefrontal cortex, posterior cingulate cortex, and lateral parietal cortex of the brain (Albert et al., 2009; Rosazza & Minati 2011) with co-activation in the ventral and dorsal medial prefrontal cortices, posterior cingulate/retrosplenial cortex, inferior parietal lobule, lateral temporal cortex, and hippocampal formation (Raichle et al., 2001). Evidence from different studies suggests that the DMN contributes to many cognitive functions, including mental exploration of social and emotional content, remembering the past (e.g., autobiographical memory), perspective taking of the belief, desire and intention (e.g., Theory of mind), and planning the future (Raichle et al., 2001; Buckner et al., 2008). DMN studies have primarily associated this network with autobiographical memory involving internally-oriented thoughts, higher-order cognitive functioning and self-referential processes (Kim, 2012; Philippi et al., 2015). Autobiographical memory is also associated with activation in MTG/STG, temporal poles, dorsal frontal, and RT cortex (Fink et al., 1996; Piefke et al., 2003). The links between DMN and auditory processing are less clear. The relationship between DMN and the right STG has not been previously reported in the hearing research literature. However, in earlier hearing-related research, differences in DMN activation have been reported in participants with and without auditory complaints. For example, a neuroimaging study of children diagnosed with APD found decreased cortical activity in DMN regions in the posterior cingulate cortex and precuneus based on ReHe analysis (Pluta et al., 2014). Pluta and colleagues (2014) suggested that dysregulation of DMN is due to failure in attention. A different study of elderly patients with tinnitus (phantom sound perception) and hearing loss also reported decreased connectivity in bilateral precuneus regions, indicating DMN dysfunction associated with internal noise/phantom noise effects (Schmidt et al., 2013). Another study involving adults with tinnitus also reported DMN disorganization in the anterior cingulate cortex and left precuneus, implying a potential role of DMN in chronic tinnitus (Chen et al., 2018).

Memory is an essential factor for cognition that is strongly linked with speech understanding in competing stimuli (e.g., noisy environment) (Hulme & Melby-Lervåg 2012). Children with APD can show deficits in short-term/working memory and attention (Moore, 2011; Sharma et al., 2014; Allen & Allan, 2014; Gokula et al., 2019). These difficulties correlate with auditory and speech processing (Hornickel et al., 2012). Our exploratory partial correlation analysis showed a significant positive correlation between PC measures and Spatial advantage skill (LiSN-S variables) in the left RT (ROI #130) (Fig. 7 A, B). The meta-analysis showed that this region is highly correlated with episodic memory, memory encoding, memory retrieval, and autobiographical memory (Fig. 7C). Studies of RT have shown its connection with the auditory cortex and memory pathways (Todd et al., 2016) and have indicated that RT plays a key role in the involvement of DMN in medial temporal regions (Andrews-Hanna et al., 2010; Kaboodvand et al., 2018). From a network neuroscience perspective, several studies have investigated PC or PC_norm_ and their relation to working memory performance during the N-back task and have suggested that the brain tends to allocate more inter-modular connectivity to fulfil the demands of this cognitive task (Shine et al., 2016; Bertolero et al., 2018; Cohen & D’Esposito, 2016; Pedersen et al., 2020). Our results for the PC/PC_norm_ analyses suggest differences between groups in inter-/intra-modular activation that could be associated with working memory differences between groups. In our study, we observed group differences for PC but not for PC_norm_. Both metrics show how a node participates in inter-modular connection, but PC_norm_ controls the influence of intra-modular connectivity (Pedersen et al., 2020). Thus, the lack of group differences in PC_norm_ suggests that the PC group differences reflect intra-modular differences in brain activity. Our hub analysis showed that the RT region functions as a connector non-hub (PC_norm_>0.5, WMZ<1) in the visual module that facilitates interactions between modules/networks. This contrasts with Kaboodvand et al.’s (2018) study of healthy adults, which suggested that RT is a provincial hub within DMN; such a difference in results could be partly related to age differences in the study populations.

Our group analysis based on Schaefer parcellation also indicated differences in the bilateral pTO (ROIs #56, #298) (Fig. 6B). Activation of bilateral Temporo-occipital cortices is associated with facial expression, visual motion, action observation, emotional expressions (Fig. 8B) and has been previously explored in reading difficulties and in facial processing difficulties in people with epilepsy (Tailby et al., 2014) and ASD (Herbert et al., 2002; Koshino et al., 2008). Although still relatively unexplored, visual processing deficits have been reported in children with APD, for example, Dawes et al.’s (2009) study of children with APD and comorbid dyslexia found visual processing difficulties as part of a multifactorial description of learning problems in these children. Furthermore, in our group comparison, we found a significant PC difference in the left IPS (ROI #93), and right posterior INS (ROI #216). The IPS region is situated in the superior parietal lobe and is thought to be highly associated with visuomotor tasks, visuospatial attention (Grefkes & Fink 2005; Silk et al., 2010; Materna et al., 2008), and working memory (Sharma et al., 2009; 2014; Gokula et al., 2019) (Fig. 8B). This outcome is consistent with reports of the co-occurrence of APD with ADHD symptoms (Chermak et al., 1998; 1999; 2002). The anterior INS is thought to be connected to a variety of human social emotions such as compassion, empathy, taste, fear, pleasure, and disgust (Lamm & Singer 2010; Mutschler et al., 2009; Ostrowsky et al. 2002; Chen et al., 2014) or homeostatic emotions such as pain (Ostrowsky et al., 2002) (Fig. 8B). However, neuroimaging studies of posterior INS have also revealed its connection to auditory cortices (Zhang et al., 2019; Dennis et al., 2014; Cloutman et al., 2012) and suggest the involvement of posterior INS with auditory processing, including allocating auditory attention, phonological processing (Bamiou et al., 2003; Remedios et al., 2009), and auditory responses observed from Heschl’s gyrus (Zhang et al., 2019).

### 4.3 Limitations and future directions

Although our research provides insights into the brain organization of children with APD, several methodological issues should be addressed in future studies.

Collecting a large sample size in a single small country like New Zealand during the global pandemic was challenging for this study. Although our sample size was, in theory, large enough to elicit reliable graph theory measures according to previous studies (Termenon et al., 2016; Andellini et al., 2015), we believe future research would benefit from data sharing approaches to gather larger data cohorts to validate our results as a generalizable finding.

We observed that the choice of brain parcellation affected our results with more significant regions in the Schaefer parcellation than the Gordon parcellation (as the number of nodes affects the interpretation of fMRI time-series and statistical comparisons). The goal of parcellation is to derive a set of homogenous brain regions with high intra-node connectivity – this is an ill-posed issue in network science that is known to affect results. This semi-arbitrary selection of parcellation and its effect on global and local properties of brain network has been discussed for a long time among researchers in the field (Zalesky et al., 2010a; Andellini et al., 2015; Termenon et al., 2016; Arslan et al., 2018; Bryce et al., 2021), and the solution that has been recommended is to apply multiple parcellation schemes to understand the influence of brain parcellation (Bryce et al., 2021). Evidence from numerous neuroimaging studies indicates that the human connectome is unique to individuals in terms of connectional traits (Mueller et al., 2013; Barch et al., 2013; Wang et al., 2015; Gordon et al., 2017), where these distinct features can be distinguished among individuals (Finn et al., 2015). Also, these unique identified patterns of brain networks could change in different tasks (Salehi et al., 2019). Since group-wise parcellations (e.g., Gordon or Schaefer parcellations) may not show these unique individual features and patterns, we hope that future research applies individualized parcellation to improve the understanding of APD’s functional organization, which would result in a deeper understanding of brain activity associated with cognitive and listening behaviors in APD.

In addition to using conventional methods where brain regions (i.e., Nodes) and their functional interactions (i.e., Edges) are the focus (Node-centric), network neuroscience has proposed a new approach for investigating the functional organization of the human brain that is centered on studying edge interactions (Edge-centric) (Faskowitz et al., 2020; 2021; Betzel, 2020; Esfahlani et al., 2021). The Edge-centric method relies on bivariate Pearson correlation (for quantifying functional connectivity) and utilizes co-fluctuations time courses between pairs of nodes instead of averaging them (Betzel, 2020). By improving temporal resolution, Edge-centric models could reveal new aspects of brain networks’ functional or structural topology (Faskowitz et al. 2020; 2021).

Another issue that arose from our cross-sectional study was that although our APD sample was diagnosed based on a comprehensive diagnostic assessment protocol (28 children diagnosed with APD), the sample was heterogeneous and included a range of children who had been recently diagnosed (39%) or diagnosed more than a year previously (60%). Some children were under treatment (32%). Future research can minimize this by utilizing a longitudinal study design to investigate changes in brain functional organization from diagnosis until treatment. Additionally, APD commonly occurs with other comorbid disorders (Sharma et al., 2009; Dawes & Bishop 2010; O’Connor, 2012; Burns, 2013; Gokula et al., 2019). Children diagnosed with APD indicate symptoms of listening difficulties when they are referred to audiology clinics, and these difficulties can be caused by auditory processing, language, cognitive, or hearing deficits (Dawes et al., 2008; Dawes & Bishop, 2010; Moore, 2012; Ankmnal-Veeranna et al., 2019; Magimairaj & Nagaraj, 2018; Dillon & Cameron, 2021). These prevalent comorbid conditions were also identified in our study populations. The participant diagnostic reports showed that almost half of the children (12 out of 28) had comorbid disorders such as dyslexia (8/29%), ADHD/ADD (3/11%), and DLD (1/3%), and the others were only diagnosed with APD (16/57%). Although we were able to see group differences in brain regions, a larger sample that separates children with APD with and without different comorbidities could better unravel differences in brain processing between these subgroups. We suggest that future studies investigate separate groups of children based on their diagnosis. This differentiation could help improve the understanding of how the functional organization of APD has been affected by comorbid disorders.

Despite a couple of reports on the structural connectivity of APD (Schmithorst et al., 2013; Farah et al., 2014), the brain structural organization of children with APD has not been fully explored. We recommend future studies by acquiring diffusion-weighted MRI data and applying network neuroscience to investigate WM microstructural organization of APD individuals to help understand neural pathways involved in listening difficulties.

## 5. CONCLUSION

In summary, this study presents new evidence of changes in brain network function in children with APD. Despite similar modular organization and whole-brain connectivity in both groups, children with APD showed atypical hub architecture compared to HC. Our results also indicated that the alteration of the brain topology in APD exists at the regional level, not at the global level. The group comparison based on the PC metric revealed significant differences in the right and bilateral auditory areas (e.g., STG) according to two methodologically different parcellations. These areas are associated with auditory processing, listening, speech, language, emotions and memory perception within DMN, indicating the role of multi-modal factors for listening difficulties in APD children. Our study underlines the importance of future research utilizing network science to understand the neural bases of APD and the use of these regional biomarkers as a potential clinical support tool.

## Data Availability

The data that support the findings of this study are available from the corresponding author upon reasonable request.

## ACKNOWLEDGEMENTS

We would like to thank all the participants and their parents for participating in this study during the COVID-19 global pandemic. We also thank Dr. Bill Keith, Mrs. Sam Marsh and Soundskill clinic in Auckland for assisting in participant recruitment. We would also like to acknowledge Dr. Catherine Morgan for her contribution in modifying MRI data acquisition protocols and Meghan van der Meer for her assistance in editing the final manuscript. Further we would like to thank the MRI technicians at CAMRI for their continued support in assisting with research studies and conducting these scans. This study was supported by Eisdell Moore Center for Hearing and Balance Research (Grant No. 3716796).

## CONFLICT OF INTEREST

The authors have no conflict of interest to declare.

## AUTHOR CONTRIBUTIONS

**Ashkan Alvand**: Conceptualization, Methodology, Investigation, Data curation, Software, Formal analysis, Visualization, Writing - original draft, Writing - review & editing. **Abin Kuruvilla-Mathew**: Conceptualization, Methodology, Investigation, Funding acquisition, Supervision, Writing - review & editing. **Ian J. Kirk**: Resources, Supervision, Writing - review & editing. **Reece R. Roberts**: Methodology, Supervision, Writing - review & editing. **Mangor Pedersen**: Methodology, Validation, Writing - review & editing. **Suzanne C. Purdy**: Conceptualization, Methodology, Project administration, Funding acquisition, Supervision, Writing - review & editing.

## SUPPORTING INFORMATION

Additional supporting information may be found online in the supporting Information section at the end of this article.

## Notes

### Competing Interest Statement

The authors have declared no competing interest.

### Author Declarations

Approval for this study was granted by The University of Auckland Human Participants Ethics committee (Date: 18/10/2019, Ref. 023546).

### Summary of Updates

Only Abstract has been revised.

