## Supporting Information for "Altered brain network topology in children with Auditory Processing Disorder: a resting-state multi-echo fMRI study" (from another version of this preprint: version 1, 2022-04-06)

1. Selection of the denoising pipeline

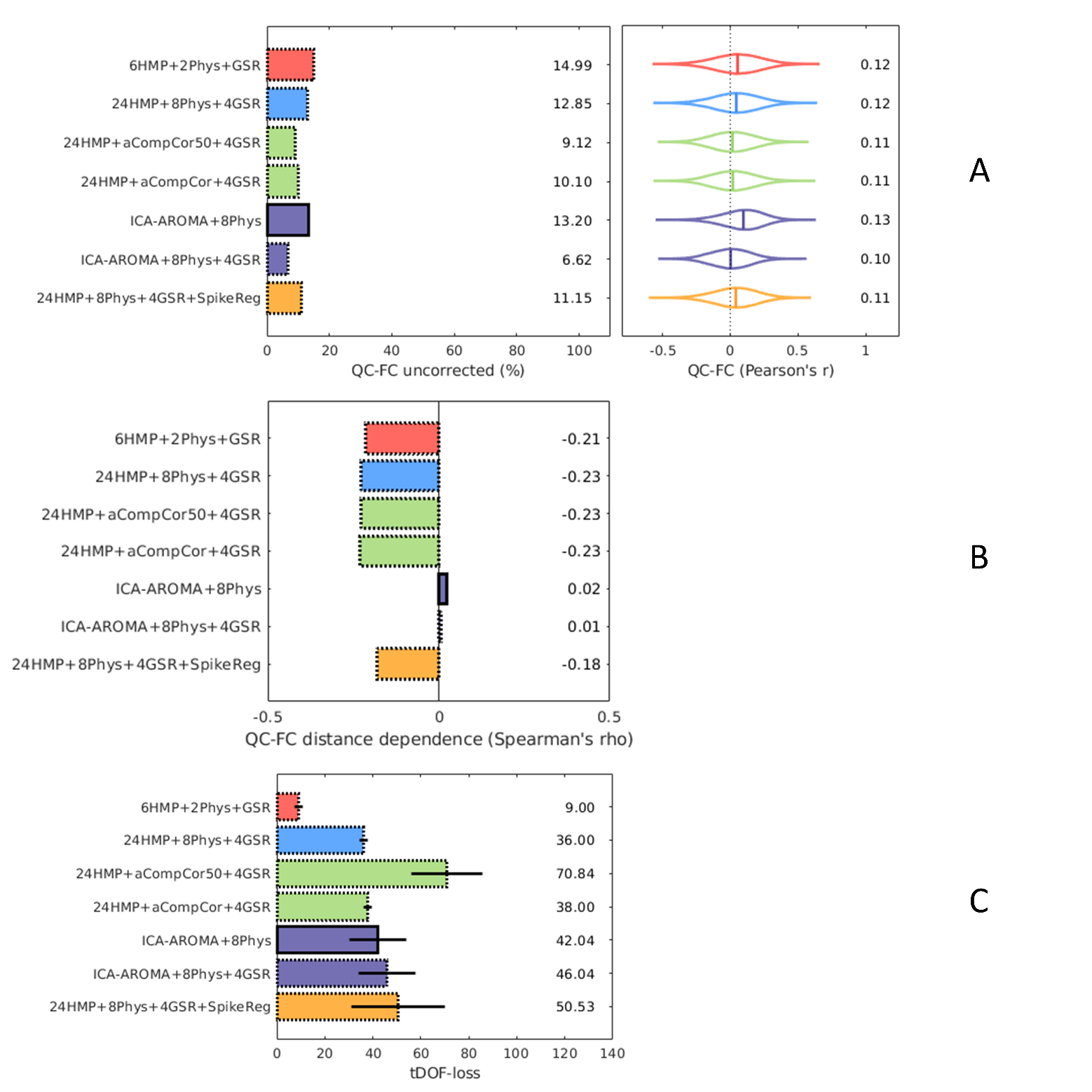

**Figure S1** Results from benchmarking the denoising pipelines. Quality control functional connectivity (QC-FC) measures were applied as recommended by Parkes and colleagues (2018) on seven different commonly used denoising procedures to quantify the efficacy (A, B; QC-FC correlation, QC-FC distance dependence) and efficiency (C; loss of temporal degree of freedom; tDOF-loss) of denoising pipelines. These pipelines were assessed based on the combination of different regressors and their temporal derivatives including head motion parameters (HMP), component-based noise correction method (aCompCor; Behzadi et al., 2007; Muschelli et al., 2014), independent component analysis strategy for automatic removal of motion artifacts (ICA-AROMA; Pruim et al., 2015) and two physiological signals (Phys) artifacts such as white matter signal (WM) and global signal regression (GSR). Based on the results the combination of ICA-AROMA+8Phys+4GSR had the best performance compared to other pipelines. See Parkes et al. (2018) and Ciric et al. (2017) for a full overview of the measure and concepts used in this benchmarking procedure.

2. Whole-brain averaged network measures

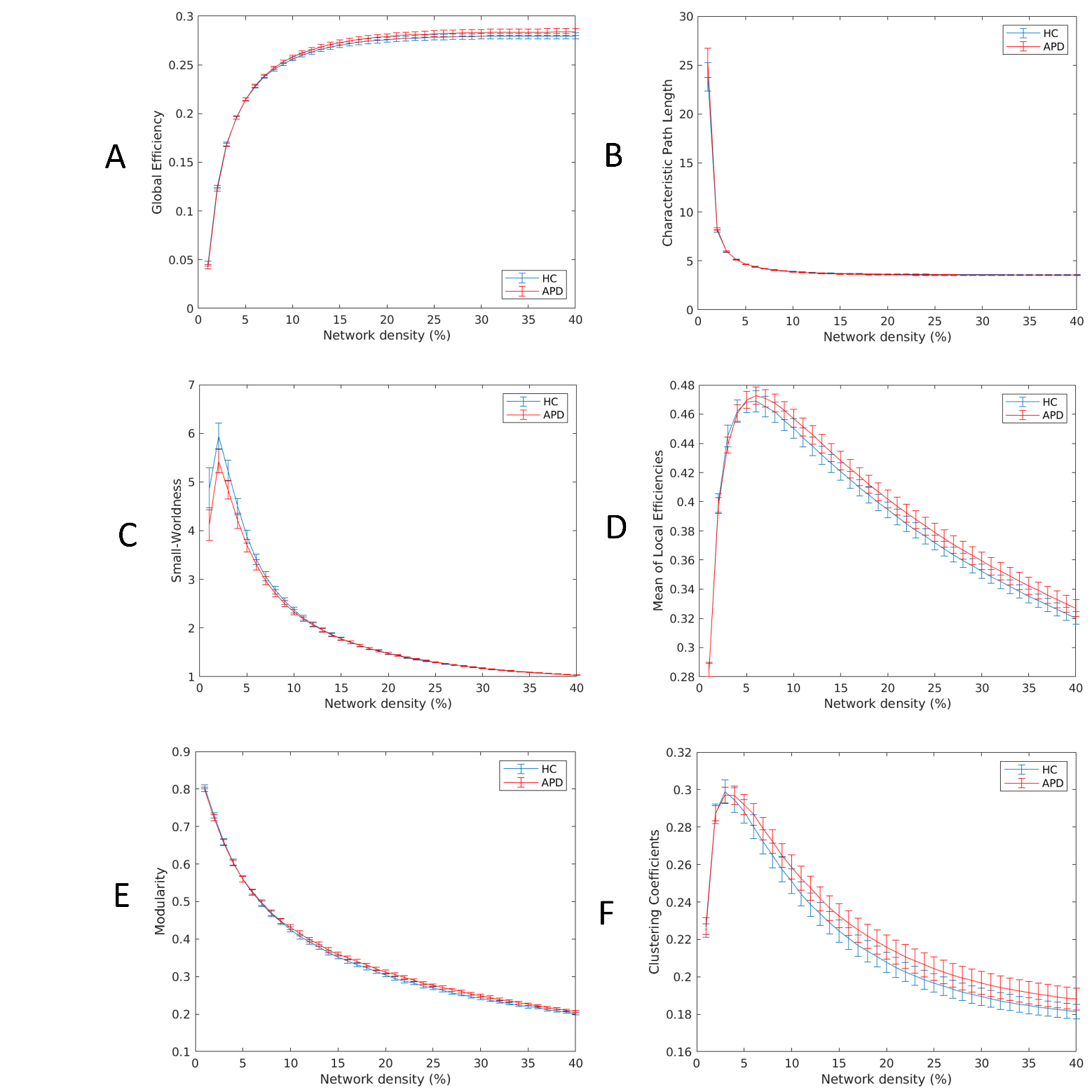

**Figure S2** Results from averaged global measure of network integration (A, global efficiency; B, characteristic path length), network segregation (D, mean local efficiency; E, modularity optimization; F, clustering coefficient,) and small-worldness metric (C). The 95% confidence interval was also calculated between APD (red line), HC (blue line) groups.

3. Hub analysis

**Table S1**

Brain hubs and their roles in APD and HC groups

| **ROIs** | **L/R** | **Regions** | **MNI** | | | **HC** | **PC_norm_** | **WMZ** | **APD** | **PC_norm_** | **WMZ** |
| --- | --- | --- | --- | --- | --- | --- | --- | --- | --- | --- | --- |
| 2 | L | SMhand | -19 | -49 | 65 |  | 0.516 | -0.061 | P | 0.421 | 1.140 |
| 5 | L | Visual | -18 | -86 | 21.6 | C | 0.537 | 1.273 | P | 0.442 | 1.213 |
| 6 | L | Default | -47 | -58 | 30.8 |  | 0.710 | 0.961 | C | 0.584 | 1.903 |
| 25 | L | Default | -6 | 42 | 35.1 | P | 0.475 | 1.856 |  | 0.560 | 0.541 |
| 27 | L | CinguloOperc | -8 | 15 | 33.8 |  | 0.520 | 0.459 | C | 0.504 | 1.037 |
| 36 | L | SMhand | -5 | -28 | 60.4 |  | 0.309 | 0.893 | P | 0.373 | 1.379 |
| 50 | L | SMhand | -29 | -45 | 61.7 |  | 0.485 | -1.135 | P | 0.344 | 1.046 |
| 52 | L | DorsalAttn | -43 | -45 | 43 | C | 0.546 | 2.512 | P | 0.486 | 1.946 |
| 57 | L | SMhand | -28 | -37 | 61.4 |  | 0.390 | 0.251 | P | 0.409 | 1.342 |
| 74 | L | DorsalAttn | -44 | 36 | 8.5 |  | 0.606 | -0.300 | P | 0.373 | 1.209 |
| 80 | L | VentralAttn | -29 | 21 | -14 | C | 0.518 | 1.508 |  | 0.602 | 0.042 |
| 82 | L | CinguloOperc | -37 | 8.9 | -0.9 | C | 0.578 | 1.042 |  | 0.557 | 0.840 |
| 87 | L | DorsalAttn | -20 | -65 | 51.4 | C | 0.567 | 1.502 |  | 0.755 | 0.571 |
| 100 | L | DorsalAttn | -46 | -58 | -7.9 | C | 0.581 | 1.466 |  | 0.649 | 0.680 |
| 103 | L | CinguloOperc | -55 | -32 | 23 | C | 0.510 | 1.224 | P | 0.498 | 1.112 |
| 111 | L | CinguloOperc | -52 | -1 | 5 | C | 0.508 | 1.915 | P | 0.477 | 1.676 |
| 126 | L | Default | -63 | -29 | -7.2 | C | 0.544 | 1.060 |  | 0.497 | 0.993 |
| 145 | L | Default | -16 | 49 | 37.2 | P | 0.475 | 1.751 |  | 0.497 | 0.943 |
| 146 | L | Default | -20 | 56 | 27.5 | C | 0.528 | 1.021 |  | 0.621 | 0.712 |
| 162 | R | Default | 12 | -52 | 34.5 |  | 0.845 | 0.968 | C | 0.740 | 1.601 |
| 163 | R | SMhand | 21 | -48 | 66.1 |  | 0.485 | -0.132 | P | 0.340 | 1.171 |
| 166 | R | Visual | 22 | -85 | 23.7 |  | 0.490 | 0.999 | P | 0.437 | 1.038 |
| 185 | R | CinguloOperc | 8.6 | 4.2 | 40.1 |  | 0.533 | 0.841 | C | 0.515 | 1.228 |
| 191 | R | SMhand | 4.8 | -27 | 64.8 |  | 0.262 | 0.943 | P | 0.379 | 1.447 |
| 195 | R | SMhand | 6.8 | -8 | 50.9 | C | 0.503 | 1.410 |  | 0.591 | 0.456 |
| 211 | R | DorsalAttn | 39 | -43 | 40.4 | P | 0.435 | 1.240 | C | 0.562 | 1.469 |
| 231 | R | VentralAttn | 49 | -27 | -0.1 |  | 0.634 | 0.520 | C | 0.638 | 1.055 |
| 245 | R | CinguloOperc | 40 | 10 | -1.6 | C | 0.635 | 1.295 |  | 0.625 | 0.830 |
| 251 | R | Visual | 27 | -55 | 54.2 | C | 0.645 | 1.094 |  | 0.754 | 0.931 |
| 252 | R | DorsalAttn | 23 | -66 | 51.8 | C | 0.595 | 1.060 |  | 0.734 | 0.429 |
| 253 | R | DorsalAttn | 32 | -64 | 33.8 | C | 0.557 | 1.049 |  | 0.637 | 0.473 |
| 269 | R | Auditory | 54 | -14 | 16.9 |  | 0.451 | 0.999 | P | 0.475 | 1.216 |
| 273 | R | FrontoParietal | 42 | 29 | 21.6 | C | 0.516 | 1.713 | P | 0.325 | 1.846 |
| 275 | R | DorsalAttn | 47 | 7.8 | 19.3 |  | 0.832 | 0.814 | C | 0.805 | 1.056 |
| 292 | R | None | 45 | 14 | -30.1 | P | 0.448 | 1.506 | C | 0.507 | 1.526 |
| 310 | R | Visual | 5.1 | -80 | 23.1 |  | 0.308 | 0.950 | P | 0.323 | 1.058 |
| 323 | R | Default | 5.9 | 55 | 29.4 | C | 0.505 | 1.287 |  | 0.455 | 0.795 |

***Note:*** Only brain regions that were different in both groups are listed in this table. Normalized participation coefficient (PC_norm_), within-module z-score degree (WMZ)**,** connector hub (C), provincial hub (P)

**Table S2**

Group difference in PC measure based on Schaefer parcellation (after control for age)

| **ROIs** | **Label** | **X** | **Y** | **Z** | ***p* value** | **t value** | **FDR** | **Bonferroni** |
| --- | --- | --- | --- | --- | --- | --- | --- | --- |
| 56 | LH_DorsAttnB_TempOcc_1 | -56 | -62 | 2 | 0.0004 | 3.4708 | 0.0375 | 0.1042 |
| 93 | LH_ContA_IPS_4 | -34 | -48 | 46 | 0.0008 | 3.2429 | 0.0413 | 0.181 |
| 124 | LH_DefaultB_Temp_3 | -52 | -20 | -8 | 0.0007 | 3.2629 | 0.0413 | 0.1727 |
| 147 | LH_TempPar_2 | -56 | -8 | -6 | 0.0003 | 3.4173 | 0.0375 | 0.1196 |
| 216 | RH_SalVentAttnA_Ins_1 | 40 | 4 | -12 | 0.0008 | 3.29 | 0.0413 | 0.1622 |
| 294 | RH_TempPar_1 | 54 | 0 | -12 | 0.0003 | 3.7096 | 0.0375 | 0.0541 |
| 298 | RH_TempPar_5 | 58 | -46 | 8 | 0.001 | 3.3764 | 0.0418 | 0.1315 |

***Note:*** TempOcc - temporalOccipital cortex, TempPar - temporal-parietal network,

Ins - intraparietal sulcus, DMN - default mode network, SalVentAttn - salience ventral attention network, Cont - control network, DorsAttn - dorsal attention network, APD - auditory processing disorder, HC - healthy control, LH - left hemisphere, RH - right hemisphere

4. Meta-analytic correlation

**Table S3**

Meta-analytic correlation between cognitive terms and significant ROIs obtained from Gordon and Schaefer parcellation

| **ROI** | **Parcellation** | **Cognitive Label** | **Meta-analytic coactivation (r)** |
| --- | --- | --- | --- |
|  | **Gordon 333** |  |  |
| 331 |  | listening | 0.42 |
|  |  | speech | 0.40 |
|  |  | acoustic | 0.36 |
|  |  | auditory | 0.36 |
|  |  | spoken | 0.36 |
|  |  | sentences | 0.36 |
|  |  | theory mind | 0.36 |
|  |  | comprehension | 0.35 |
|  |  | linguistic | 0.35 |
|  |  | social | 0.35 |
|  |  | speaker | 0.35 |
|  |  | voice | 0.34 |
|  |  | sounds | 0.33 |
|  |  | mind | 0.32 |
|  |  | autobiographical | 0.30 |
|  |  | mental states | 0.30 |
|  |  | speech perception | 0.28 |
|  |  | language | 0.27 |
|  |  | music | 0.26 |
|  |  | vocal | 0.25 |
|  | **Schaefer 300** |  |  |
| 294 |  | listening | 0.39 |
|  |  | theory mind | 0.39 |
|  |  | speech | 0.37 |
|  |  | social | 0.37 |
|  |  | sentences | 0.35 |
|  |  | mind | 0.35 |
|  |  | comprehension | 0.34 |
|  |  | spoken | 0.34 |
|  |  | autobiographical | 0.34 |
|  |  | linguistic | 0.33 |
|  |  | speaker | 0.33 |
|  |  | acoustic | 0.32 |
|  |  | auditory | 0.32 |
|  |  | voice | 0.32 |
|  |  | sounds | 0.3 |
|  |  | mentalizing | 0.28 |
|  |  | language | 0.26 |
|  |  | music | 0.24 |
|  |  | junction | 0.24 |
|  |  | vocal | 0.23 |
| 56 |  | motion | 0.37 |
|  |  | actions | 0.32 |
|  |  | perception | 0.31 |
|  |  | object | 0.25 |
|  |  | gestures | 0.25 |
|  |  | language | 0.25 |
|  |  | visual motion | 0.25 |
|  |  | action observation | 0.24 |
|  |  | lexical | 0.23 |
|  |  | linguistic | 0.23 |
|  |  | verbs | 0.23 |
|  |  | sentence | 0.22 |
|  |  | semantic | 0.22 |
|  |  | words | 0.22 |
|  |  | auditory | 0.21 |
|  |  | comprehension | 0.21 |
|  |  | spoken | 0.21 |
|  |  | visual | 0.20 |
|  |  | audiovisual | 0.20 |
|  |  | listening | 0.20 |
| 93 |  | tasks | 0.67 |
|  |  | working memory | 0.59 |
|  |  | demands | 0.43 |
|  |  | calculation | 0.41 |
|  |  | attention | 0.39 |
|  |  | memory wm | 0.38 |
|  |  | arithmetic | 0.32 |
|  |  | visuospatial | 0.31 |
|  |  | execution | 0.29 |
|  |  | interference | 0.28 |
|  |  | symbolic | 0.28 |
|  |  | maintenance | 0.28 |
|  |  | spatial attention | 0.28 |
|  |  | memory load | 0.26 |
|  |  | phonological | 0.26 |
|  |  | eye movements | 0.26 |
|  |  | task difficulty | 0.26 |
|  |  | action | 0.25 |
|  |  | performance | 0.25 |
|  |  | target | 0.24 |
| 124 |  | comprehension | 0.45 |
|  |  | sentences | 0.45 |
|  |  | language | 0.44 |
|  |  | linguistic | 0.43 |
|  |  | listening | 0.40 |
|  |  | spoken | 0.39 |
|  |  | speech | 0.38 |
|  |  | semantic | 0.37 |
|  |  | auditory | 0.35 |
|  |  | syntactic | 0.35 |
|  |  | speaker | 0.34 |
|  |  | sounds | 0.32 |
|  |  | words | 0.31 |
|  |  | language comprehension | 0.30 |
|  |  | lexical | 0.30 |
|  |  | acoustic | 0.29 |
|  |  | phonological | 0.28 |
|  |  | voice | 0.28 |
|  |  | audiovisual | 0.27 |
|  |  | speech perception | 0.26 |
| 147 |  | listening | 0.73 |
|  |  | spoken | 0.71 |
|  |  | speech | 0.7 |
|  |  | auditory | 0.64 |
|  |  | sounds | 0.63 |
|  |  | linguistic | 0.62 |
|  |  | acoustic | 0.61 |
|  |  | sentences | 0.6 |
|  |  | comprehension | 0.58 |
|  |  | language | 0.58 |
|  |  | voice | 0.56 |
|  |  | music | 0.48 |
|  |  | syntactic | 0.44 |
|  |  | hearing | 0.44 |
|  |  | pitch | 0.42 |
|  |  | lexical | 0.41 |
|  |  | semantic | 0.4 |
|  |  | words | 0.4 |
|  |  | vocal | 0.4 |
|  |  | phonological | 0.39 |
| 216 |  | pleasant | 0.28 |
|  |  | neutral | 0.28 |
|  |  | taste | 0.27 |
|  |  | unpleasant | 0.27 |
|  |  | disgust | 0.27 |
|  |  | pain | 0.26 |
|  |  | emotional | 0.25 |
|  |  | affective | 0.24 |
|  |  | fear | 0.23 |
|  |  | aversive | 0.22 |
|  |  | anxiety | 0.20 |
|  |  | happy | 0.19 |
|  |  | reactivity | 0.19 |
|  |  | valence | 0.19 |
|  |  | arousal | 0.18 |
|  |  | facial expressions | 0.18 |
|  |  | intensity | 0.17 |
|  |  | angry | 0.17 |
|  |  | emotional faces | 0.17 |
|  |  | ptsd | 0.17 |
| 298 |  | facial expression | 0.34 |
|  |  | motion | 0.33 |
|  |  | perception | 0.32 |
|  |  | junction | 0.25 |
|  |  | audiovisual | 0.25 |
|  |  | action observation | 0.22 |
|  |  | gaze | 0.2 |
|  |  | listening | 0.2 |
|  |  | language | 0.18 |
|  |  | linguistic | 0.18 |
|  |  | modality | 0.18 |
|  |  | action | 0.17 |
|  |  | auditory | 0.17 |
|  |  | auditory visual | 0.17 |
|  |  | speech | 0.17 |
|  |  | sentences | 0.16 |
|  |  | spoken | 0.16 |
|  |  | voice | 0.15 |
|  |  | acoustic | 0.14 |
|  |  | social cognition | 0.14 |

***Note:*** The top 20 associated cognitive terms are shown for each significant ROIs according to Gordon and Schaefer parcellations. The meta-analytic terms were derived from the Neurosynth database (Yarkoni et al., 2011). Cognitive terms were selected based on their meta-analytic coactivation scores (Pearson correlation; uncorrected)
